# Cohort Profile: The Mendelian Randomization in Pregnancy (MR-PREG) collaboration - Improving evidence for prevention and treatment of adverse pregnancy and perinatal outcomes

**DOI:** 10.1101/2025.03.22.25324447

**Authors:** Nancy McBride, Gemma L Clayton, Ana Goncalves Soares, Qian Yang, Tom A Bond, Amy Taylor, Charikleia Chatzigeorgiou, Elisabeth Aiton, Jane West, Maria C Magnus, Deborah A Lawlor, Maria Carolina Borges

## Abstract

**Purpose:** Adverse pregnancy and perinatal outcomes (APPOs), including pre-term birth, pre-eclampsia, and gestational diabetes, can result in maternal and neonatal morbidity and mortality, parental anxiety, and increased health care costs. Better understanding of causes of APPOs is essential to inform lifestyle and pharmaceutical interventions for their prevention and management. Given the difficulty of undertaking randomised control trials in pregnant women, triangulating evidence from across different methods with different sources of bias could improve our understanding of the causes of APPOs. The purpose of the Mendelian Randomization in Pregnancy (MR-PREG) collaboration is to support triangulation of evidence from genetic (e.g., Mendelian randomization [MR]) and non-genetic (e.g., partner negative controls) methods to explore causal effects of maternal exposures on a comprehensive set of APPOs

**Participants:** The MR-PREG collaboration includes individual participant data from three birth cohorts (two from the UK and one from Norway) and UK Biobank, and summary data from FinnGen and publicly available genome wide association studies (GWAS). We have harmonised data across studies so that currently includes exposures on up to 34 APPOs in up to 678,001 women.

**Findings to date:** The main aims of MR-PREG are to improve the evidence base for 1) prevention, by advancing our understanding of maternal modifiable causes of APPOs, 2) better understanding of the effect of pre-existing conditions on APPOs, and 3) treatment, by advancing our knowledge of the efficacy and safety of existing medications that women may require for pre-existing conditions, and identifying and testing the efficacy and safety of novel medications, and those that might be repurposed to effectively and safely treat APPOs. To date, our published research mainly addresses aims 1 and 3; some examples include triangulation of evidence from MR, conventional multivariable regression and a paternal negative control, showing that higher maternal body mass index increases the risk of many APPOs, and identification of maternal circulating metabolites and proteins that may influence birthweight.

**Future Plans:** Our future priorities include increasing diversity in the MR-PREG collaboration by expanding participant representation from non-European ancestries. We are also integrating molecular data, such as circulating protein levels and placental transcriptomics, to better understand the molecular mechanisms underlying APPOs. Additionally, we are using exome and whole-genome sequencing to identify novel causal genes for APPOs and advance our knowledge on candidate targets for APPOs.

**Strengths and limitations:**

- We have curated data for 34 APPOs and harmonized data across multiple studies to support large-scale investigations of causes of APPOs.
- The scope and type of the data supports triangulation of evidence from a range of genetic and non-genetic methods, with different unrelated sources of bias, to identify causes of APPOs.
- Over the coming year we will enhance the data to enable identification of molecular mechanism underlying APPOs.
- MR-PREG has limited power to detect causal effects on rarer APPOs, such as congenital anomalies and low Apgar scores at 5 minutes, particularly when using genetic methods. Future work will include larger samples and rare genetic variant data.
- Participants are mostly of European ancestry; future efforts will be focussed on diversifying the ancestry representation in the data.

## Introduction

A substantial proportion of pregnant women experience adverse pregnancy or perinatal outcomes (APPOs), such as miscarriage, stillbirth, gestational diabetes mellitus (GDM), gestational hypertension (GH), pre-eclampsia (PE), pre-term birth (PTB), or having a small (SGA) or large for gestational age (LGA) baby. Globally, one in six recognised pregnancies results in miscarriage ^1^, one in six women develops GDM ^2^, one in ten women has hypertension during pregnancy ^3^, and one in ten babies is born preterm ^4^. Some APPOs are causes of severe morbidity and mortality for mothers and their babies ^5^. For illustration, hypertensive disorders of pregnancy (HDP), including GH and PE, account for approximately 14% of all maternal deaths worldwide ^6^. In addition, APPOs are associated with long-term adverse physical and mental health outcomes, and high costs to families, healthcare systems and society ^7^ ^8^. In the UK, the short-term costs of miscarriage alone are estimated to be around £471 million per year due to costs to health services, families and loss of productivity ^1^.

The marked variation in the prevalence of some APPOs across regions and over time, although influenced by different screening and diagnostic practices, indicates that they are preventable^2^. Better understanding the underlying causes of APPOs is crucial for guiding effective interventions to prevent them. This knowledge would also enable antenatal care services to provide women and couples with accurate advice on the most plausible risk factors for reducing APPOs, potentially reducing the often conflicting advice given to women about exposures that could impact their and their babies’ health during pregnancy. Furthermore, APPOs are often related, meaning that preventing one may help reduce the burden of others. For example, both GDM and HDP can influence fetal growth, leading to the delivery of LGA and SGA babies, respectively ^9^ ^10^. Therefore, preventing GDM and HDP is a way of avoiding fetal over/under growth and the related complications.

There is also an urgent need to better understand the effects of pharmaceutical treatments during pregnancy, both for preventing and managing APPOs and for treating pre-existing conditions that are increasingly common among women of reproductive age, such as autoimmune, mental health, thyroid, reproductive, and cardiometabolic disorders ^11-14^. An increasing number of women start pregnancy on medication, and the use of medication among pregnant women has also been rising over the past few decades ^15^. Despite that, pregnant women are rarely included in pre-licensure randomized controlled trials (RCTs) due to the concerns about potential teratogenic effects of drugs and the potential impact of drug-dosing on the physiological changes of pregnancy ^16^ ^17^. As a result, evidence on the benefits and risks of drugs for mother and baby is poor, investment in new drug development for APPOs is scarce, and very few medications are explicitly licensed for use during pregnancy. This leaves mothers and their babies exposed to unknown risks of drugs, compels pregnant women and their doctors to undertreat medical conditions, and forces them to make difficult decisions about which medications to prescribe/maintain and at which dose. It also means that APPOs, such as GDM and GH, are managed less well than equivalent conditions outside of pregnancy (i.e., type 2 diabetes and hypertension) ^16^ ^17^.

RCTs are the gold standard method for testing the impact of lifestyle and pharmaceutical interventions to prevent or treat APPOs. For several years, national bodies have highlighted the need for clinical trials of medications in pregnant women, largely to no avail ^16^ ^18^. Acknowledging this, the UK 2021 Report of the Commission on Human Medicines Expert Working Group on Optimising Data On Medicines Used During Pregnancy recommended i) the use of routine healthcare data in research, and ii) the use of observational research to investigate safety of medicines used in pregnancy and during breastfeeding should be commissioned and a system of ongoing surveillance should be established and funded ^18^.

In the absence of well-powered, well-conducted RCTs, we need to make the best use of observational data to improve the current evidence base on causes of APPOs and medication efficacy and safety during pregnancy. More than 20 years ago, the use of genetic variants to infer causal effects of exposures − a method known as Mendelian randomization (MR) − was first proposed ^19^. MR leverages the random allocation of genetic variants at conception to investigate the effects of modifiable risk factors on health outcomes. This method mitigates confounding by socioeconomic, behavioural, or health-related factors that frequently affect traditional observational study analyses (**Figure 1**) ^19-21^. Since then, its application has expanded significantly, including for assessing the effects of a few maternal risk factors on a small range of APPOs ^22^ ^23^ ^24^.

**Figure 1.**
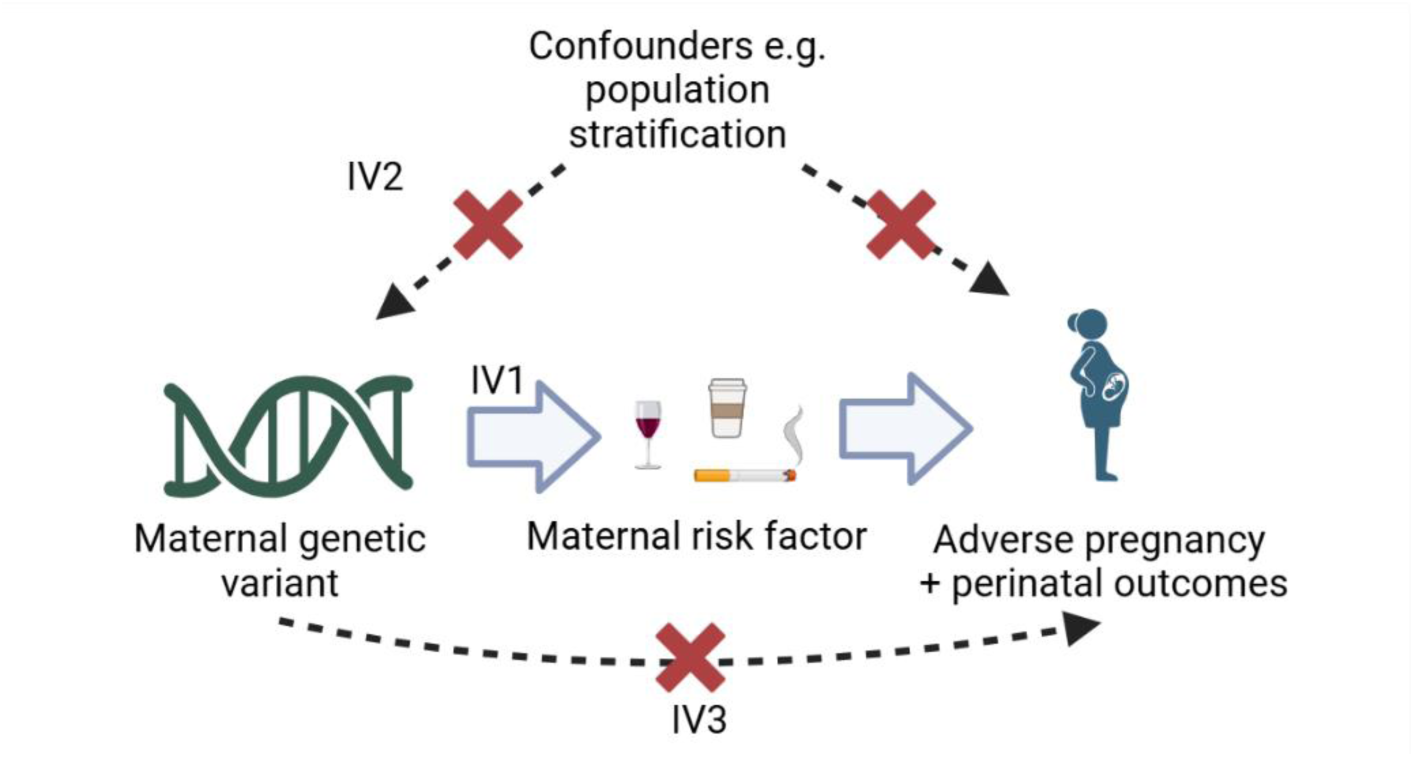
Summary of the Mendelian randomisation assumptions to estimate the presence of an effect of maternal risk factors on adverse pregnancy and perinatal outcomes.

### Assumptions

- The maternal genetic variant(s) are statistically robustly associated with the maternal risk factor during pregnancy **[relevance assumption, IV1]**
- There is no confounding of the maternal genetic variant(s) and adverse pregnancy and perinatal outcomes (APPOs) (i.e., population level confounders such as population structure, assortative mating, and intergenerational effects) ^25^ **[independence assumption, IV2]**
- The maternal genetic variant is not associated with APPOs other than through its association with the maternal risk factor **[exclusion restriction criteria, IV3]**

In addition, MR is increasingly used for assessing the effects of medications outside of pregnancy (aka drug target MR). The validity of drug target MR is supported by proof-of-concept studies comparing MR findings to RCT results for established medications, such as antihypertensives and statins ^26^. Drug targets validated by genetic evidence have higher rates of success and MR is increasingly used to prioritise (or de-prioritise) new drug targets to be tested in RCTs ^27-29^.

As with all methods, MR is limited by violation of its assumptions. Triangulation of evidence acknowledges, and exploits the fact that all methods have sources of bias ^30^. It involves integrating multiple lines of evidence using one or more different approaches (e.g., different analytical methods, different data-sources, or different study designs) with different and unrelated key sources of bias. If results are consistent, despite the different sources of bias, this increases the credibility of that being the correct causal effect (whether it suggests a protective, detrimental or no effect), as it would be unlikely for different biases to produce different results. Where there is disagreement across the different methods, prior specification of key sources of bias, the direction of these biases, and statistical efficiency for each can help inform whether the inconsistency is explained by the different biases or power for each approach, which can inform what further sensitivity analyses and/or other approaches are needed to obtain a valid causal estimate.

The Mendelian Randomization in Pregnancy (MR-PREG) collaboration was established to improve causal understanding of the effects of maternal lifestyle and health factors on APPOs, as well as efficacy and safety of medication use in pregnancy. In this paper, we provide the aims of the MR-PREG collaboration, describe the characteristics of the contributing studies, our methods for data generation and exploring causal effects, as well as summarising findings to date and discussing ongoing and future work.

**Aim 1:** Using triangulation of evidence to improve knowledge on the impact of maternal lifestyle factors on APPOs

The MR-PREG collaboration uses triangulation of genetic methods (e.g., MR, co-localisation, rare variant, and joint rare and common variant analyses) and non-genetic methods (e.g., conventional multivariable regression, and negative paternal controls) to explore the effect of a range of maternal modifiable lifestyle factors, such as smoking, alcohol intake, coffee consumption, adiposity, sleeping habits, and physical activity, on the risk of multiple APPOs (**Figure 2**) (**Supplementary Table 1**) ^30^.

**Figure 2.**
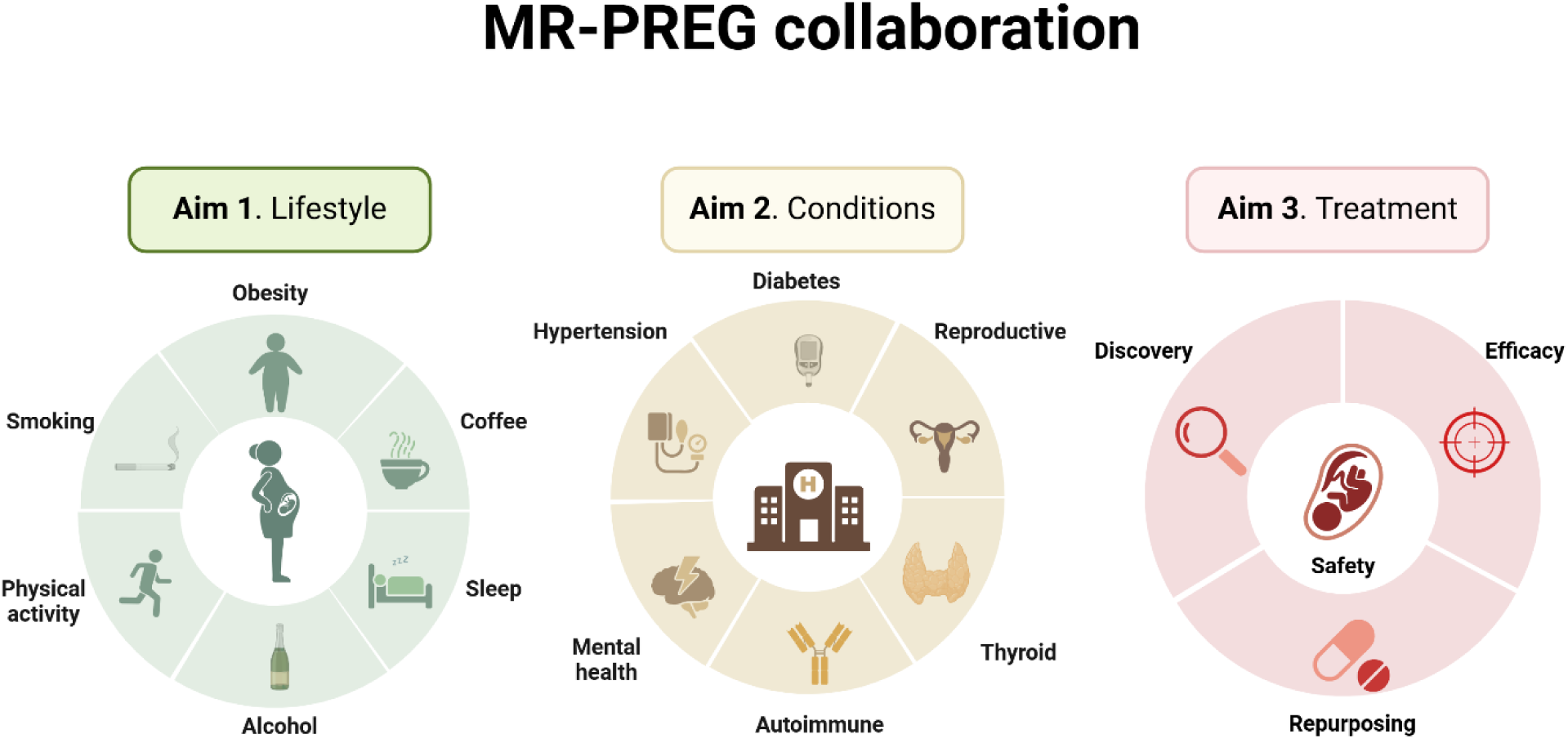
MR-PREG collaboration aims.

**Aim 2:** Better understanding the effect of maternal predisposition to conditions, such as autoimmune, cardiovascular, hormonal, musculoskeletal, and mental health conditions, on APPOs An increasing number of women begin pregnancy with one or more pre-existing conditions that may elevate the risk of multiple APPOs ^12^. A systematic understanding of the overall impact of these conditions on APPO risk is important for guiding clinical decision-making in the management and treatment of such conditions during pregnancy. Within the MR-PREG collaboration, we employ MR to investigate how genetic predisposition to common conditions in women of reproductive age - such as autoimmune, cardiovascular, hormonal, musculoskeletal, and mental health conditions - affects APPO risk (**Figure 2**).

**Aim 3:** Leveraging genomics to explore drug efficacy and safety of medications in pregnancy

The MR-PREG collaboration aims to improve the evidence on the benefits and risks of medications during pregnancy using drug target MR. We investigate potential effects of medications for mothers and their babies by investigating the efficacy and safety in pregnancy of medications used to treat pre-existing conditions, discovering new candidate drug targets to prevent/treat APPOs, and identifying opportunities to repurpose existing drugs, designed to treat conditions unrelated to pregnancy, to prevent/treat APPOs (**Figure 2**).

## Cohort description

### Adverse pregnancy and perinatal outcomes (APPOs)

A key focus of the MR-PREG collaboration is on assessing the causal effect of exposures on multiple outcomes simultaneously, so that we have a more complete picture of the potential beneficial, detrimental, or null effect of an exposure on APPOs. This ‘outcome-wide epidemiology’ approach ^31^ is informative so that women, their partners and healthcare providers have a more balanced view of the potential adverse and beneficial effects of risk factors than what might be obtained from studies that focus on a single or small number of outcomes.

We have included 34 binary/categorical outcomes that can occur in pregnancy, delivery or the first year after birth, and 2 underlying continuous traits in MR-PREG. **Table 1** shows the APPOs and underlying traits available in MR-PREG together with the number of women who have phenotypic and genetic data on each of these and the number of cases for the outcomes. Cohort-specific APPOs definitions and exclusions are provided in supplementary material (**Supplementary Tables 2A** and **2B)**, and as well as sample size for binary/categorical outcomes (**Supplementary Table 3A** and **3B f**or maternal phenotype and genetic data, respectively, and **Supplementary Table 3C** for offspring genetic data), and mean and standard deviations for continuous traits (**Supplementary Table 3D**). **Supplementary Tables 4A** and **4B** describe any deviations from the collaboration definition for each study.

**Table 1.** Adverse pregnancy and perinatal outcomes (APPOs) and underlying traits, total available sample and respective number of cases.

| Type | Outcomes/Traits | Total | Cases N (%) |
| --- | --- | --- | --- |
| Pregnancy | Hypertensive disorders of pregnancy | 541,768 | 32,549 (6.0%) |
|  | Gestational hypertension | 527,932 | 20,777 (3.9%) |
|  | Pre-eclampsia | 678,001 | 19,566 (2.9%) |
|  | Gestational diabetes | 934,566 | 26,346 (2.8%) |
|  | Depression | 147,601 | 22,785 (15.4%) |
|  | Anaemia | 86,167 | 4,105 (4.8%) |
|  | Pregnancy loss | 298,868 | 94,152 (31.5%) |
|  | Miscarriage | 486,217 | 89,086 (18.3%) |
|  | Sporadic miscarriage | 241,159 | 55,888 (23.2%) |
|  | Recurrent miscarriage | 321,756 | 6,233 (1.9%) |
|  | Stillbirth | 207,670 | 6,331 (3.0%) |
|  | Hyperemesis | 478,466 | 4,235 (0.9%) |
|  | Severity of nausea and vomiting | 82,978 |  |
|  | Nausea only |  | 7,166 (8.6%) |
|  | Nausea and vomiting |  | 30,133 (36.3%) |
|  | On medication |  | 31,210 (37.6%) |
| Delivery | Induction of labour | 95,239 | 14,495 (15.2%) |
|  | Premature rupture of membranes | 310,621 | 21,839 (7.0%) |
|  | Caesarean section | 233,969 | 33,909 (14.5%) |
|  | Emergency C-section | 90,438 | 9,165 (10.1%) |
|  | Elective C-section | 87,283 | 6,004 (6.9%) |
|  | Very pre-term birth | 85,274 | 1,214 (1.4%) |
|  | Pre-term birth | 518,849 | 29,987 (5.8%) |
|  | Post-term birth | 425,673 | 27,841 (6.5%) |
|  | Low birth weight | 275,214 | 18,617 (6.8%) |
|  | High birth weight | 266,835 | 6,679 (2.5%) |
|  | Small for gestational age | 89,033 | 6,882 (7.7%) |
|  | Large for gestational age | 96,305 | 10,468 (10.9%) |
|  | Low Apgar score at 1 minute | 86,668 | 4,950 (5.7%) |
|  | Low Apgar score at 5 minutes | 78,739 | 876 (1.1%) |
| Postnatal | NICU admission | 77,285 | 6,996 (9.1%) |
|  | Congenital anomalies | 74,701 | 3,569 (4.8%) |
|  | Congenital heart disease | 74,701 | 612 (0.8%) |
|  | Breastfeeding initiation | 84,668 | 66,408 (78.4%) |
|  | Breastfeeding established | 81,258 | 58,035 (71.4%) |
|  | Breastfeeding sustained | 65,189 | 44,401 (68.1%) |
|  | Breastfeeding duration | 82,429 |  |
|  | 1 to 3 |  | 6,372 (7.7%) |
|  | 4 to 6 |  | 49,123 (59.6%) |
|  | >6 |  | 3,419 (4.1%) |
| Underlying traits | Birth weight* | 289,846 | - |
|  | Gestational age* | 226,810 | - |

\* continuous traits

### Data sources

Currently, the MR-PREG collaboration includes data from four core studies, which comprise three prospective birth cohorts [the Avon Longitudinal Study of Parents and Children (ALSPAC), Born in Bradford (BiB), and the Norwegian Mother, Father and Child Cohort Study (MoBa)], and a biobank [UK Biobank (UKB)]. Data from additional sources are also used, such as from publicly available biobanks (FinnGen) and genome-wide association studies (GWAS) meta-analyses, described below.

### Core studies

A brief description of each of the core studies is presented below and the participant characteristics are summarised in **Table 2**. **Supplementary Figures 1-4** show the flow of participants from recruitment into their cohorts to inclusion in our analyses. The APPOs each study contributed to MR-PREG are summarised in **Supplementary Tables 2A and 2B**. Details on genotyping in each study are presented in supplementary material.

**Table 2.** Maternal participant characteristics in the participating core studies^#^.

| Characteristics | ALSPAC | BiB | MoBa | UK Biobank |
| --- | --- | --- | --- | --- |
| Total N | 13,895 | 9,407 | 93,512 | 224,423 |
| Age, mean (SD) | 28.0 (5.0) | 27.4 (5.6) | 30.1 (4.7) | 25.3 (4.6)* |
| Missing | 369 (2.7%) | 1,169 (12.4%) | 458 (0.5%) | 45,402 (20.2%) |
| Ethnicity (White), N (%) | 11,600 (97.4%) | 3,430 (36.4%) | 88,989 (95.1%) | 211,677 (94.6%) |
| Missing | 1,982 (14.3%) | 1,025 (10.1%) | 2,021 (2.1%) | 706 (0.3%) |
| Education (university level), N (%) | 1,544 (12.9%) | 1,432 (11.8%) | 50,047 (53.5%) | 65,334 (29.5%)** |
| Missing | 1,890 (13.6%) | 1,186 (12.6%) | 11,064 (11.8%) | 2,714 (1.2%) |
| Index of multiple deprivation, (N%) |  |  |  |  |
| 1st quintile (most deprived) | 1,628 (13.9%) | 5,232 (55.6%) | NA | 44,857 (20.0%)<sup>§</sup> |
| 5th quintile (least deprived) | 3,045 (26.0%) | 158 (1.7%) | NA | 44,805 (20.0%) |
| Missing | 2,197 (15.8%) | 1,171 (12.4%) |  | 384 (0.2%) |
| Parity (nulliparous), N(%) | 5,619 (44.7%) | 3,768 (40.0%) | 42,135 (45.1%) | 179,385 (83.6%) |
| Missing | 1,333 (9.6%) | 345 (3.6%) | 399 (0.4%) | 9,762 (4.3%) |
| Smoking during pregnancy (ever), N (%) | 3,301 (25.9%) | 1,408 (14.9%) | 5,939 (6.4%) | NA |
| Missing | 1,124 (8.1%) | 1,185 (12.6%) | 16,824 (18.0%) |  |
| Alcohol intake during pregnancy, N (%) | 8,036 (64.0%) | 1,707 (18.1%) | 2,472 (2.6%) | NA |
| Missing | 1,340 (9.6%) | 2,114 (22.9%) | 27,648 (29.6%) |  |
| Body mass index, mean (SD) | 22.9 (3.8) | 26.1 (5.7) | 24.1 (4.3) | 27.1 (5.1)** |
| Missing | 2,711 (19.5%) | 1,562 (16.6%) | 11,561 (12.3%) | 888 (0.4%) |

<sup>#</sup> Participants included if they had data on at least one APPO
\* age at first birth; \*\* information assessed at recruitment and not at pregnancy; <sup>§</sup> this corresponds to deprivation index and quintiles were constructed internally (not based on national comparisons) Abbreviations: ALSPAC, The Avon Longitudinal Study of Parents and Children; BiB, Born in Bradford; MoBa, The Norwegian Mother, Father and Child cohort study; SD, standard deviation; NA, information not available in this study. This includes all available data at the outcome level.

#### ALSPAC: The Avon Longitudinal Study of Parents and Children

The Avon Longitudinal Study of Parents and Children (ALSPAC) is a prospective birth cohort that started recruiting pregnant women resident in the former county of Avon (centred around the city of Bristol), with delivery dates between April 1991 and December 1992. A total of 14,541 women (ALSPAC-G0) were enrolled during pregnancy (14,676 fetuses) and gave birth to 14,062 live children (ALSPAC-G1) ^32^ ^33^. Women responded to four questionnaires during pregnancy (average 8, 12, 18, and 32 weeks gestation) and two postpartum (average 8 weeks and 8 months). Biological samples were collected during pregnancy (blood and urine) and birth (cord blood and placenta). Maternal anthropometrics was based on self-report collected at 12 weeks gestation and children had anthropometrics measured at birth. Obstetric records were also linked to the participants. Genetic data is available for mothers, partners and children (details on genotyping array and imputation are presented in supplementary material). Children of the children (ALSPAC-G2) are also being assessed and followed-up, but this data has not yet been included in MR-PREG. The study website contains details of all the data available through a fully searchable data dictionary and variable search tool: http://www.bristol.ac.uk/alspac/researchers/our-data/. Ethical approval for the study was obtained from the ALSPAC Ethics and Law Committee and the Local Research Ethics Committees. Full details of the ALSPAC consent procedures are available on the study website (http://www.bristol.ac.uk/alspac/researchers/research-ethics/http://www.bristol.ac.uk/alspac/researchers/research-ethics/).

#### BiB: Born in Bradford

Born in Bradford (BiB) is a prospective birth cohort that recruited women with expected delivery dates between March 2007 and December 2010. Most women were recruited at their oral glucose tolerance test (OGTT) at approximately 26–28 weeks’ gestation, which was offered to all women booked for delivery at Bradford Royal Infirmary, except those with known diabetes, during the recruitment period. In BiB, most of the obstetric population consists of women of White British or Pakistani origin (together accounting for 81%, with the remaining women being of other ancestries). A total of 12,453 women (13,776 pregnancies) were enrolled during pregnancy who gave birth to 13,858 live children. Women had anthropometrics measured at recruitment and responded to questionnaires during pregnancy and postpartum to provide information about breastfeeding > 6 months. Biological samples were collected during pregnancy (blood and urine) and birth (cord blood) and women consented to routine primary and secondary care data linkage. Full details of the study methodology were reported previously ^34^. Genetic data is available for mothers and children (details on genotyping array and imputation presented in supplementary material). Ethical approval for the study was granted by the Bradford National Health Service Research Ethics Committee (ref 06/Q1202/48). The study website provides further cohort details and an overview of available data (https://borninbradford.nhs.uk/).

#### MoBa: The Norwegian Mother, Father and Child Cohort Study

The Norwegian Mother, Father and Child Cohort Study (MoBa) is a prospective birth cohort that recruited pregnant women from all over Norway from 1999-2008. The cohort includes approximately 95,200 mothers, 75,200 fathers, and 114,500 children. Mothers and their partners responded to questionnaires during pregnancy (15, 22, and 30 weeks gestation) and postpartum (6 months). Blood samples were obtained from both parents during pregnancy and from mothers and children (cord blood) at birth. Maternal anthropometrics were collected from a questionnaire at 15 weeks gestation. Data was linked to the Medical Birth Registry (MBRN), which is a national health registry containing information about all births in Norway. The current study is based on version 12 of the quality-assured data files released for research in 2019. Genetic data is available for mothers, children and partners (details on genotyping array and imputation presented in supplementary material). The establishment of MoBa and initial data collection was based on a license from the Norwegian Data Protection Agency and approval from The Regional Committees for Medical and Health Research Ethics. The MoBa cohort is currently regulated by the Norwegian Health Registry Act. Ethical approval for our study was obtained from The Regional Committees for Medical and Health Research Ethics (ref 2018/1256).

#### UKB: UK Biobank

UK Biobank (UKB) is an adult cohort that retrospectively collected relevant data on APPOs. All people in the UK National Health Service (NHS) registry aged between 40-69 years and living within an approximately 25-mile radius from one of the 22 study centres were invited to participate in UKB between 2006-2010 ^35^ ^36^. A total of 500,000 adults (5.5% of the ∼9.2 million invited) were recruited into the study (54.4% females). Information was assessed at baseline via a self-completed questionnaire, physical measures (including anthropometrics), and collection of non-fasting blood, urine, and saliva. Participants have been followed up by linkage to electronic health records and a subset of participants responded to online questionnaires. Hospital Episode Statistics (HES) from 1997 for England, 1998 for Wales and 1981 for Scotland are available ^37^. HES data also contain maternity-related admissions for England and Wales. It is important to note that not every participant has a hospital inpatient record, as not all have been admitted to hospital within the period covered. Genetic data from UKB participants is available (details on genotyping array and imputation presented in supplementary material). Ethical approval for UKB was obtained from the Northwest Multi-Centre Research Ethics Committee (MREC), and MR-PREG collaboration studies are linked to UKB application number 23938. The UKB showcase website contains details of the data available: https://biobank.ndph.ox.ac.uk/showcase/.

#### Additional data sources

To increase statistical power for MR analyses, the MR-PREG collaboration also uses genetic association data for APPOs from publicly available datasets – i.e., FinnGen and several publicly available GWAS meta-analyses – as described below.

#### FinnGen

FinnGen is the nationwide network of Finnish biobanks, which are linked to national electronic registries that provide information on prescriptions and diseases (ICD9-10 codes). FinnGen includes data from 500,348 individuals (282,064 females and 218,284 males) [12^th^ data release (R12)]. Besides clinical endpoints, which also include data on some APPOs, genetic data is also available. The APPOs and number of cases and controls for which FinnGen contributed are detailed in **Supplementary Table 3B**. The Coordinating Ethics Committee of the Helsinki and Uusimaa Hospital District has approved the FinnGen consortium (Nr HUS/990/2017). More information about FinnGen can be found on the website https://www.finngen.fi/en. The metadata from FinnGen used by the MR-PREG collaboration is publicly available at https://www.finngen.fi/en/access_results.

#### GWAS meta-analyses

At the time of writing, the MR-PREG collaboration has harmonised and quality-controlled data from GWAS meta-analyses for GDM [5,485 cases and 347,856 controls from the GENetics of Diabetes in Pregnancy Consortium (GenDIP) consortium ^38^]; PE [9,515 cases and 157,719 controls from the International Pregnancy Genetics (InterPregGen) consortium ^39^]; gestational duration-related traits [18,797 PTB cases and 260,246 controls, 15,972 post-term birth cases and 115,307 controls, and 195,555 individuals with gestational age at delivery from the Early Growth Genetics (EGG) consortium ^40^]; post-natal depression [17,339 cases and 53,426 controls from the Psychiatric Genomics consortium (PGC) ^41^]. Details are provided in **Supplementary Table 5**, including the number of participants, outcome definition and source of data (e.g., electronic health records, maternal report, research data collection), and availability of maternal and/or fetal genetic effects.

### Genetic association data for adverse pregnancy and perinatal outcomes

For studies with access to individual-level data (ALSPAC, BiB, MoBa, and UKB), we conducted GWAS analyses to generate genetic association data for APPOs, enabling two-sample MR analyses across the aims of the MR-PREG collaboration. The procedures used for conducting GWAS and quality control in each study are described in detail in the **Supplementary text**. We excluded genetic variants with low imputation accuracy (INFO score < 0.4) and/or low minor allele frequency (MAF < 0.01). In addition, we excluded studies that overlap with public GWAS metanalyses or that contributed with less than 50 cases for a given APPO and applied genomic control to each study. We then pooled study-specific genetic association data on APPOs using inverse variance weighted fixed-effects meta-analyses implemented in METAL (v. 2020-05-05) ^42^ and estimated the Cochrane’s Q statistics to explore between-study heterogeneity. Separate meta-analyses were conducted for maternal and offspring genetic effects. We used GWASInspector to evaluate the quality of GWAS summary data from each GWAS and for the meta-analyses ^43^.

#### Conditional estimates

The MR-PREG collaboration is mostly interested in the causal effects of maternal exposures on APPOs. Due to the correlation between maternal and offspring genetics, accounting for offspring genetic effects is crucial to reliably interpret MR findings testing maternal exposure effects (**Box 1**, **Figure 3a** and **Figure 3b**). Genetically instrumented maternal exposure effects (not biased due to exclusion restriction violation via fetal effects) and genetically instrumented fetal effects (not confounded by maternal genotype) can be estimated. To account for mutually adjusted maternal and fetal genetic effects we used a weighted linear model (WLM) implemented using DONUTS (Decomposing nature and nurture using GWAS summary statistics) software ^44^ to estimate mutually adjusted maternal and fetal genetic effects at each SNP. The WLM calculates conditional genetic effects (i.e., the mutually adjusted coefficients for maternal, offspring and paternal genotype, fitted jointly in the same model) as linear combinations of the marginal genetic effects (i.e., the coefficients for maternal, offspring and paternal genotype, estimated separately in potentially overlapping samples). In MoBa, because we also had genotype data on a large sample of fathers, we did the same to account for paternal genetic effects. Details can be found in **Supplementary text.**

**Figure 3a.**
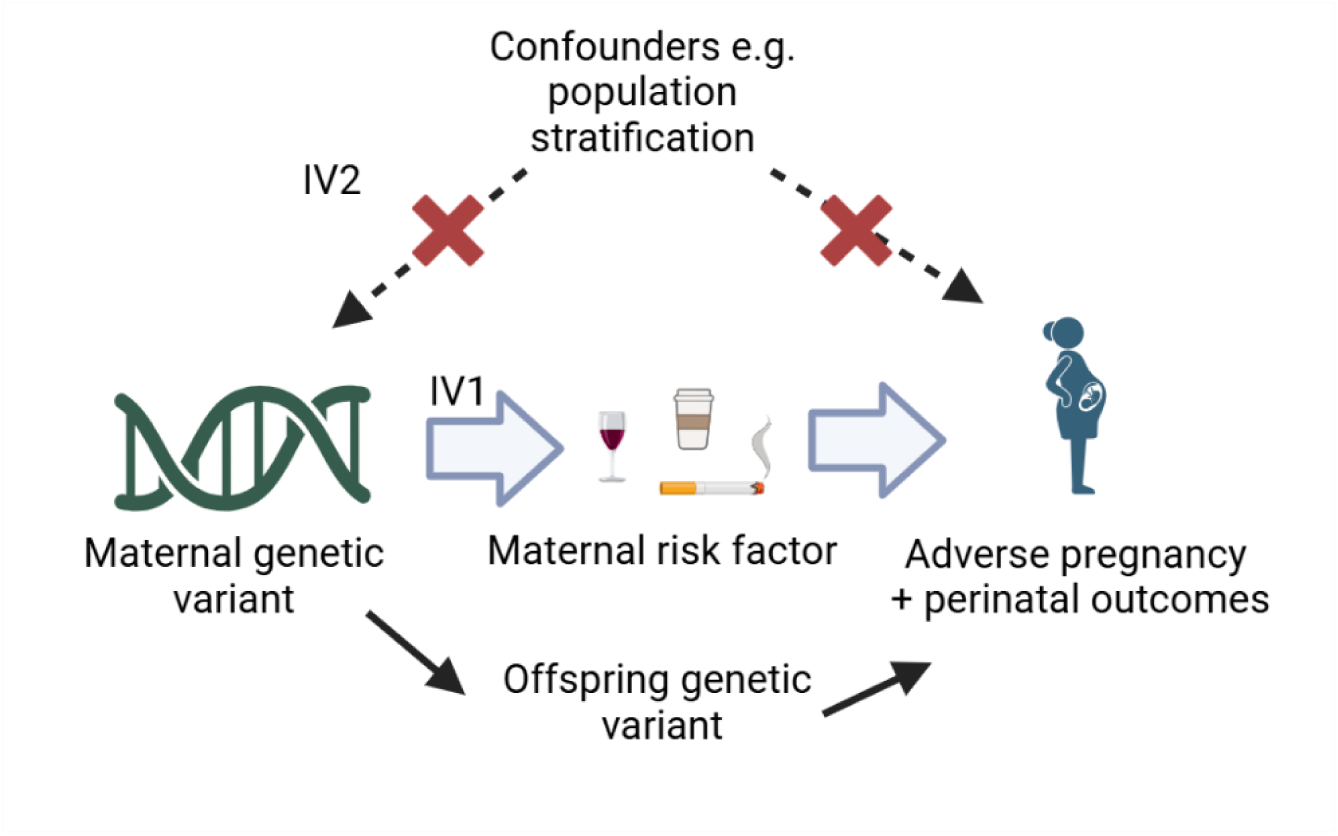
Illustration of how the exclusion restriction assumption may be violated.

**Figure 3b.**
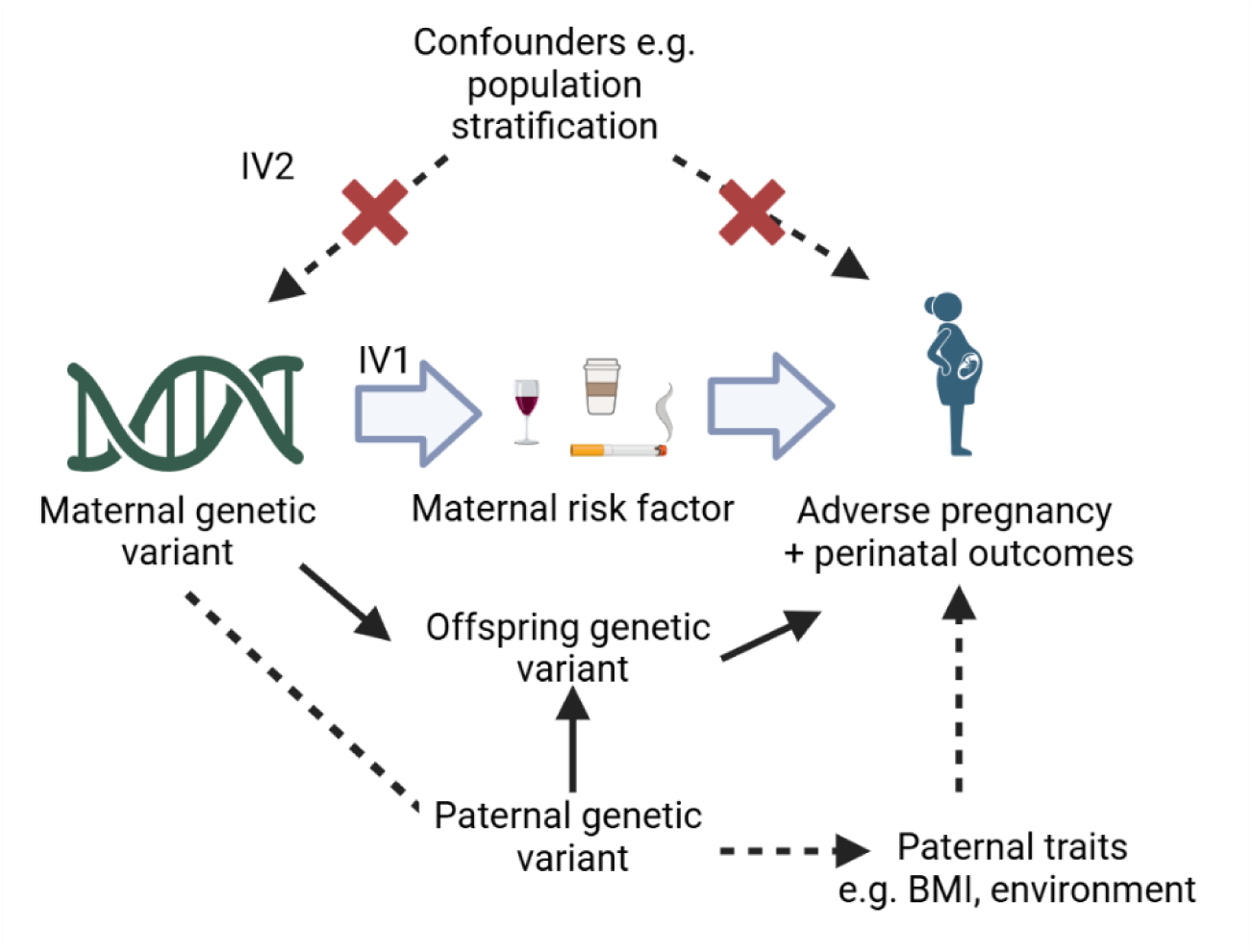
Illustration of how collider bias might be induced in MR analysis.

##### Box 1

Approaches to check the Mendelian randomisation assumptions (**Figure 1**), specific to maternal risk factors and adverse pregnancy and perinatal outcomes (APPOs). These are further sensitivity methods specific to this scenario, beyond standard MR sensitivity methods such as weighted median and MR-Egger etc.

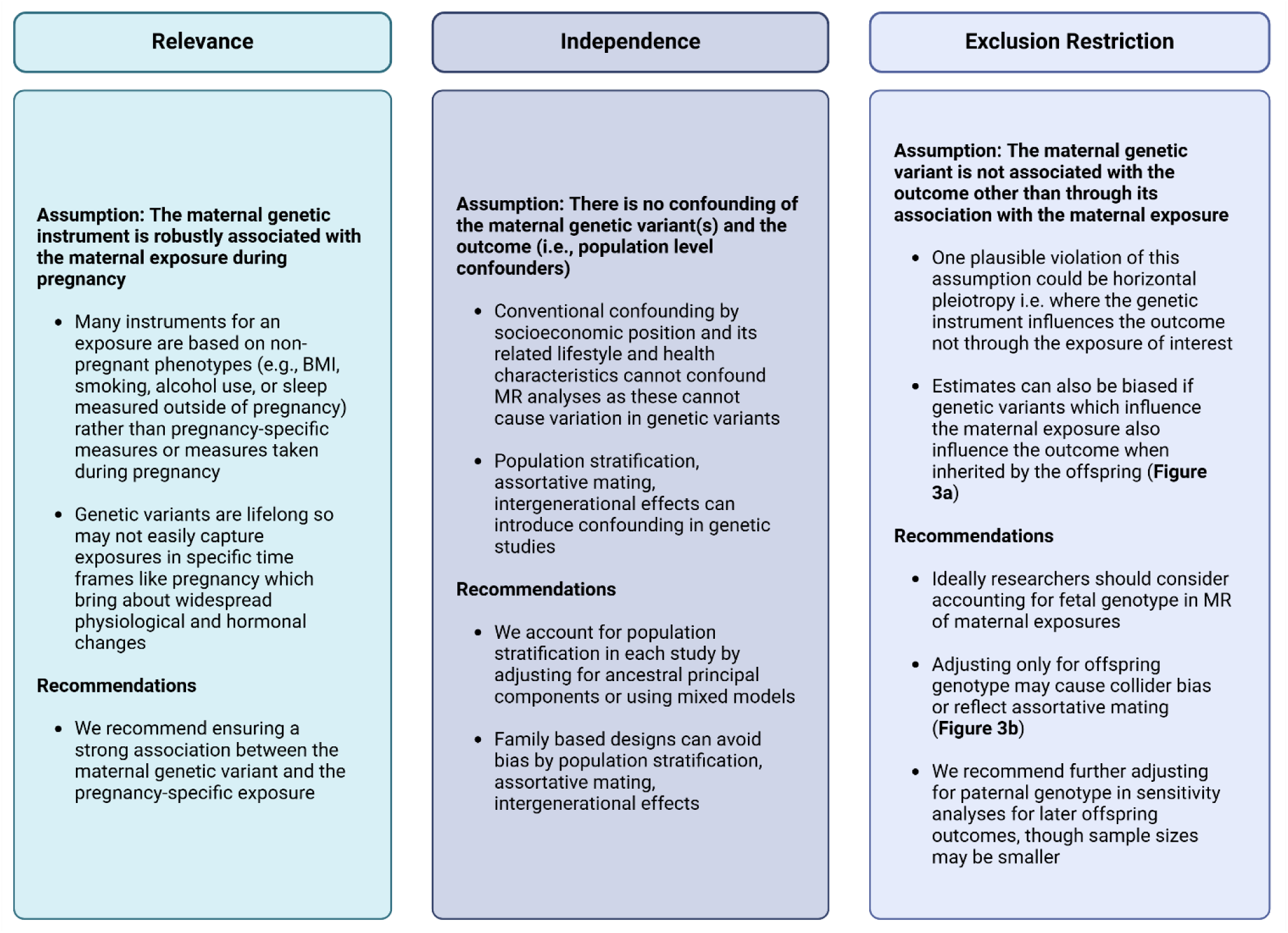

Adjusting for offspring genetic variants only (which are a collider variable) (**Figure 3b**) means that a false association between maternal and paternal genetic variants can be created (shown as a dashed line). Such associations can also arise via assortative mating. This will only cause bias if the outcome is influenced by paternal genetic variants, independently of maternal and offspring genetic variants. However, by additionally adjusting for paternal genetic variants, the pathway highlighted in black dashed arrows can be closed.

## Findings to date

To date there have been 8 peer-reviewed publications ^23^ ^45-52^ and 1 preprint ^53^ resulting from the MR-PREG collaboration. We briefly describe a selection of these here to give readers an indication of the different ways in which MR-PREG can be used to explore effects of exposures on APPOs.

Most publications to date align with **Aim 1**, enhancing our understanding of how maternal lifestyle factors influence the risk of APPOs, thereby informing interventions aimed at their prevention. For example, by triangulating evidence from MR, conventional multivariable regression and paternal negative control, we have shown that higher maternal BMI increases the risk of GH, PE, GDM, pre-labour membrane rupture, induction of labour, Caesarean section, LGA, high birthweight, low Apgar score, and admission to a neonatal intensive care unit, and reduced risk of breastfeeding. We also found evidence of higher maternal BMI reducing the odds of SGA and having no effect on perinatal depression ^49^. Triangulating evidence from MR and conventional multivariable regression, we found evidence that insomnia may increase the risk of perinatal depression but does not appear to influence most other APPOs that we were able to explore ^51^. In a study focussed on fetal growth trajectories, assessed by repeat ultrasound scan measures in two cohorts, we triangulated evidence from MR, conventional multivariable regression and paternal negative control study and found evidence of a consistent linear dose-response association of maternal smoking with fetal growth from early in the second trimester onwards. No major growth deficit was found in women who quit smoking early in pregnancy ^54^.

In relation to **Aim 3**, we have published preliminary work identifying novel molecular targets (i.e., proteins and metabolites) causally related to APPOs. In one study, we used MR to investigate the effect of 1,139 maternal and fetal circulating proteins on offspring birthweight. We found evidence that higher maternal levels of PCSK1 potentially increase birthweight whilst higher maternal levels of LGALS4 potentially decrease birthweight. Conversely, higher fetal levels of PCSK1 potentially decrease birthweight and of LGALS4 potentially increase birthweight. Higher fetal LEPR increased birthweight. Results support maternal and fetal protein effects on birthweight, implicating roles for glucose metabolism, energy balance, and vascular function ^53^. We have also identified maternal circulating metabolites beyond glucose that may influence birthweight, such as amino acids like glutamine ^46^. In further studies, we have used conventional multivariable regression and MR to explore the potential effect of > 1,000 maternal circulating mass spectrometry metabolites on the risk of offspring congenital heart disease. We found that pregnancy amino acid metabolism, androgenic steroid lipids, and levels of succinylcarnitine could be important contributing factors for congenital heart disease (CHD) ^50^.

## Strengths and limitations

The collaboration has created curated data for 34 APPOs and harmonised data across several studies to support large-scale investigations of causes of APPOs. In addition, the generation of genetic association data enables well-powered genetic studies, including MR studies, to improve causal knowledge of many key targets for lifestyle and pharmaceutical interventions aimed at preventing APPOs. Data on equivalent exposures in partners and on a wide range of plausible confounders, enables triangulation across genetic and non-genetic approaches, and also appropriate sensitivity analyses. In relation to those sensitivity analyses the collaboration has also derived conditional estimates so that maternal genetic variants can be used as an instrument to test the effect of maternal exposures without biases related to offspring genetic effects.

Despite the size and scale of the data, we are still underpowered to detect causal effects on rare APPOs, such as congenital anomalies and low Apgar scores. Furthermore, participants are of predominantly European ancestry, except for BiB. Enhancing the ancestry diversity in the data contributing to the collaboration is a key priority as outlined below in ‘Collaborations and future plans’. Finally, despite our best efforts to derive accurate and standardised APPO definitions, there is inevitably a considerable degree of misclassification, especially where some studies derived their data only from self-reported information or only from hospital electronic health records.

## Collaborations and future plans

We are currently focussing on systematically assessing the effects of predisposition to autoimmune, mental health, thyroid, reproductive, and cardiometabolic disorders on APPOs (**Aim 2**), discovering new candidate drug targets for APPOs (**Aim 3**), and investigating the efficacy, safety and potential of repurposing existing drugs during pregnancy (**Aim 3**).

A future priority is to increase diversity in the genetic background of MR-PREG collaboration participants, given the current effort is predominantly focussed on participants of European ancestry. This will enable us to improve the internal validity of the MR-PREG studies (e.g., selecting better genetic instruments through using trans-ancestral data to improve statistical fine mapping) but also external validity (e.g., testing transportability of effects across ancestries). Additionally, we are incorporating more molecular data into MR-PREG studies to better understand the molecular mechanisms underlying APPOs, for example, placental transcriptomics and proteomics.

Finally, to enhance our work identifying molecular targets for the prevention and treatment of APPOs (**Aim 3**), we are utilizing exome and whole-genome sequencing data to pinpoint causal genes linked to these adverse outcomes.

We are advancing collaborations with additional large-scale studies including genetic and APPO data, with a focus on rarer outcomes such as congenital anomalies and participants of non-European ancestry to increase statistical power for key safety outcomes and ancestry diversity. Genetic association data for APPOs generated by the collaboration can only be used for research that is covered by data agreements with current contributing studies.

## Data availability

The ALSPAC access policy that describes the proposal process in detail including any costs associated with conducting research at ALSPAC, which may be updated from time to time and is available at: https://www.bristol.ac.uk/medialibrary/sites/alspac/documents/researchers/dataaccess/ALSPAC_Access_Policy.pd

Data is available upon request from Born in Bradford: https://borninbradford.nhs.uk/research/how-to-access-data/

Data from MoBa are available from the Norwegian Institute of Public Health after application to the MoBa Scientific Management Group (see its website https://www.fhi.no/en/op/data-access-from-health-registries-health-studies-and-biobanks/data-access/applying-for-access-to-data/ for details).

Researchers can apply for access to the UK Biobank data via the Access Management System (AMS) (https://www.ukbiobank.ac.uk/enable-your-research/apply-for-access).

## Author contributions

The MR PREG collaboration was conceptualised by DAL and MCB. GC, NM, AGS, MCB, QY, TB, CC, EA, AT undertook data curation and generated GWAS pipelines for the individual cohorts. QY and MCB ran the meta-analysis. TB and MCB generated the WLM models. GC, NM, MCB, AGS and DAL, wrote, and all authors edited and approved the manuscript.

## Acknowledgements

The authors would like to thank Professor Dave Evans, Dr Marwa Al-Arab, Dr Alba Fernandez-Sanles, Dr Alice Carter, Dr Helena Urquijo, Jevvy Huang, and Peiyuan Huang for their assistance with statistical analysis, GWAS, and technical support.

## Funding declaration

All cohort specific funding information is detailed in the **Supplementary text**.

This publication is the work of the authors, who will serve as guarantors for the contents of this paper.

DAL, MCB, GC, AGS, TB, QY, AT, HC, EA, PH, and NM work in a unit supported by the University of Bristol and UK Medical Research Council (MC_UU_00032/5) and the British Heart Foundation (AA/18/1/34219). AGS is supported by the European Union’s Horizon 2020 research and innovation programme (grant agreement No 874739, LongITools). AGS, GC and DAL are supported by the European Union’s Horizon Europe Research and Innovation Programme (grant agreement No 101137146, STAGE, via UKRI grant number 10099041). EA is supported by a Wellcome Trust PhD studentship (228276/Z/23/Z). MCM is supported by the Research Council of Norway through its Centres of Excellence funding scheme (project No 262700) and the European Research Council under the European Union’s Horizon 2020 research and innovation program (ERC Starting Grant, INFERTILITY grant agreement No 947684).

## References

1. Quenby S, Gallos ID, Dhillon-Smith RK, et al. Miscarriage matters: the epidemiological, physical, psychological, and economic costs of early pregnancy loss. The Lancet 2021;397(10285):1658–67. doi: 10.1016/S0140-6736(21)00682-6

2. Wang H, Li N, Chivese T, et al. IDF Diabetes Atlas: Estimation of Global and Regional Gestational Diabetes Mellitus Prevalence for 2021 by International Association of Diabetes in Pregnancy Study Group’s Criteria. Diabetes Res Clin Pract 2022;183:109050. doi: 10.1016/j.diabres.2021.109050 [published Online First: 20211206]

3. Abalos E, Cuesta C, Grosso AL, et al. Global and regional estimates of preeclampsia and eclampsia: a systematic review. Eur J Obstet Gynecol Reprod Biol 2013;170(1):1–7. doi: 10.1016/j.ejogrb.2013.05.005 [published Online First: 20130607]

4. Ohuma EO, Moller AB, Bradley E, et al. National, regional, and global estimates of preterm birth in 2020, with trends from 2010: a systematic analysis. Lancet 2023;402(10409):1261–71. doi: 10.1016/s0140-6736(23)00878-4

5. Souza JP, Day LT, Rezende-Gomes AC, et al. A global analysis of the determinants of maternal health and transitions in maternal mortality. The Lancet Global Health 2024;12(2):e306–e16. doi: 10.1016/S2214-109X(23)00468-0

6. Geller SE, Koch AR, Garland CE, et al. A global view of severe maternal morbidity: moving beyond maternal mortality. Reproductive Health 2018;15(1):98. doi: 10.1186/s12978-018-0527-2

7. Catov JM, Margerison-Zilko C. Pregnancy as a window to future health: short-term costs and consequences. Am J Obstet Gynecol 2016;215(4):406–7. doi: 10.1016/j.ajog.2016.06.060

8. Rich-Edwards JW, Fraser A, Lawlor DA, et al. Pregnancy Characteristics and Women’s Future Cardiovascular Health: An Underused Opportunity to Improve Women’s Health? Epidemiologic Reviews 2014;36(1):57–70. doi: 10.1093/epirev/mxt006

9. Brito Nunes C, Borges MC, Freathy RM, et al. Understanding the Genetic Landscape of Gestational Diabetes: Insights into the Causes and Consequences of Elevated Glucose Levels in Pregnancy. Metabolites 2024;14(9) doi: 10.3390/metabo14090508 [published Online First: 20240920]

10. Wu P, Green M, Myers JE. Hypertensive disorders of pregnancy. BMJ 2023;381:e071653. doi: 10.1136/bmj-2022-071653

11. Angum F, Khan T, Kaler J, et al. The Prevalence of Autoimmune Disorders in Women: A Narrative Review. Cureus 2020;12(5):e8094. doi: 10.7759/cureus.8094 [published Online First: 20200513]

12. Singh M, Wambua S, Lee SI, et al. Autoimmune diseases and adverse pregnancy outcomes: an umbrella review. BMC Medicine 2024;22(1):94. doi: 10.1186/s12916-024-03309-y

13. Molenaar NM, Bais B, Lambregtse-van den Berg MP, et al. The international prevalence of antidepressant use before, during, and after pregnancy: A systematic review and meta-analysis of timing, type of prescriptions and geographical variability. J Affect Disord 2020;264:82–89. doi: 10.1016/j.jad.2019.12.014 [published Online First: 20191209]

14. Conrad N, Molenberghs G, Verbeke G, et al. Trends in cardiovascular disease incidence among 22 million people in the UK over 20 years: population based study. Bmj 2024;385:e078523. doi: 10.1136/bmj-2023-078523 [published Online First: 20240626]

15. Subramanian A, Azcoaga-Lorenzo A, Anand A, et al. Polypharmacy during pregnancy and associated risk factors: a retrospective analysis of 577 medication exposures among 1.5 million pregnancies in the UK, 2000-2019. BMC Medicine 2023;21(1):21. doi: 10.1186/s12916-022-02722-5

16. Sciences AoM. Understanding pregnancy: Accelerating the development of new therapies for pregnancy specific conditions. In: Foundation BHPatC, ed., 2023.

17. National Academies Sciences E, Medicine Consensus Study Report;. Advancing Clinical Research with Pregnant and Lactating Populations: Overcoming Real and Perceived Liability Risks, 2024.

18. UK Commission on Human Medicines. Report of the Commission on Human Medicines Expert Working Group on Optimising Data on Medicines used During Pregnancy, 2021.

19. Smith GD, Ebrahim S. ’Mendelian randomization’: can genetic epidemiology contribute to understanding environmental determinants of disease? Int J Epidemiol 2003;32(1):1–22. doi: 10.1093/ije/dyg070

20. Lawlor DA, Harbord RM, Sterne JA, et al. Mendelian randomization: using genes as instruments for making causal inferences in epidemiology. Stat Med 2008;27(8):1133–63. doi: 10.1002/sim.3034

21. Zheng J, Baird D, Borges MC, et al. Recent Developments in Mendelian Randomization Studies. Curr Epidemiol Rep 2017;4(4):330–45. doi: 10.1007/s40471-017-0128-6 [published Online First: 20171122]

22. Yan M, Chen Z, Tang J, et al. Association between gestational diabetes mellitus and offspring health: a two-sample mendelian randomization study. BMC Pregnancy and Childbirth 2025;25(1):321. doi: 10.1186/s12884-025-07423-4

23. Taylor K, Wootton RE, Yang Q, et al. The effect of maternal BMI, smoking and alcohol on congenital heart diseases: a Mendelian randomisation study. BMC Medicine 2023;21(1):35. doi: 10.1186/s12916-023-02731-y

24. Tyrrell J, Richmond RC, Palmer TM, et al. Genetic Evidence for Causal Relationships Between Maternal Obesity-Related Traits and Birth Weight. (1538-3598 (Electronic))

25. Morris TT, Davies NM, Hemani G, et al. Population phenomena inflate genetic associations of complex social traits. Science Advances 2020;6(16):eaay0328. doi: 10.1126/sciadv.aay0328

26. Schmidt AF, Finan C, Gordillo-Marañón M, et al. Genetic drug target validation using Mendelian randomisation. Nature Communications 2020;11(1):3255. doi: 10.1038/s41467-020-16969-0

27. Holmes MV, Richardson TG, Ference BA, et al. Integrating genomics with biomarkers and therapeutic targets to invigorate cardiovascular drug development. Nat Rev Cardiol 2021;18(6):435–53. doi: 10.1038/s41569-020-00493-1 [published Online First: 20210311]

28. Zheng J, Haberland V, Baird D, et al. Phenome-wide Mendelian randomization mapping the influence of the plasma proteome on complex diseases. Nature Genetics 2020;52(10):1122–31. doi: 10.1038/s41588-020-0682-6

29. Atia A, Aboeldahab H, Wageeh A, et al. Safety and Efficacy of Proprotein Convertase Subtilisin-Kexin Type 9 Inhibitors After Acute Coronary Syndrome: A Meta-analysis of Randomized Controlled Trials. American Journal of Cardiovascular Drugs 2024;24(1):83–102. doi: 10.1007/s40256-023-00621-5

30. Lawlor DA, Tilling K, Davey Smith G. Triangulation in aetiological epidemiology. Int J Epidemiol 2016;45(6):1866–86. doi: 10.1093/ije/dyw314

31. VanderWeele TJ. Outcome-wide Epidemiology. Epidemiology 2017;28(3):399-402. doi: 10.1097/ede.0000000000000641

32. Boyd A, Golding J, Macleod J, et al. Cohort Profile: the ‘children of the 90s’ – the index offspring of the Avon Longitudinal Study of Parents and Children. Int J Epidemiol 2013;42 doi: 10.1093/ije/dys064

33. Fraser A, Macdonald-Wallis C, Tilling K, et al. Cohort profile: The Avon Longitudinal Study of Parents and Children: ALSPAC mothers cohort. Int J Epidemiol 2013;42 doi: 10.1093/ije/dys066

34. Wright J, Small N, Raynor P, et al. Cohort Profile: the Born in Bradford multi-ethnic family cohort study. Int J Epidemiol 2013;42(4):978–91. doi: 10.1093/ije/dys112 [published Online First: 20121012]

35. Littlejohns TJ, Sudlow C, Allen NE, et al. UK Biobank: opportunities for cardiovascular research. Eur Heart J 2019;40(14):1158–66. doi: 10.1093/eurheartj/ehx254

36. Sudlow C, Gallacher J, Allen N, et al. UK biobank: an open access resource for identifying the causes of a wide range of complex diseases of middle and old age. PLoS Med 2015;12(3):e1001779. doi: 10.1371/journal.pmed.1001779 [published Online First: 20150331]

37. UK Biobank. UK Biobank hospital inpatient data, 2023.

38. Pervjakova N, Moen GA-O, Borges MC, et al. Multi-ancestry genome-wide association study of gestational diabetes mellitus highlights genetic links with type 2 diabetes. (1460-2083 (Electronic))

39. Steinthorsdottir V, McGinnis R, Williams NO, et al. Genetic predisposition to hypertension is associated with preeclampsia in European and Central Asian women. Nat Commun 2020;11(1):5976. doi: 10.1038/s41467-020-19733-6 [published Online First: 20201125]

40. Solé-Navais P, Flatley C, Steinthorsdottir V, et al. Genetic effects on the timing of parturition and links to fetal birth weight. Nat Genet 2023;55(4):559–67. doi: 10.1038/s41588-023-01343-9 [published Online First: 20230403]

41. Guintivano J, Byrne EM, Kiewa J, et al. Meta-Analyses of Genome-Wide Association Studies for Postpartum Depression. Am J Psychiatry 2023;180(12):884–95. doi: 10.1176/appi.ajp.20230053 [published Online First: 20231018]

42. Willer CJ, Li Y, Abecasis GR. METAL: fast and efficient meta-analysis of genomewide association scans. Bioinformatics 2010;26(17):2190–1. doi: 10.1093/bioinformatics/btq340 [published Online First: 20100708]

43. Ani A, van der Most PJ, Snieder H, et al. GWASinspector: comprehensive quality control of genome-wide association study results. Bioinformatics 2021;37(1):129–30. doi: 10.1093/bioinformatics/btaa1084

44. Wu Y, Zhong X, Lin Y, et al. Estimating genetic nurture with summary statistics of multigenerational genome-wide association studies. Proceedings of the National Academy of Sciences 2021;118(25):e2023184118. doi: doi:10.1073/pnas.2023184118

45. Yang Q, Magnus MC, Kilpi F, et al. Evaluating causal associations of chronotype with pregnancy and perinatal outcomes and its interactions with insomnia and sleep duration: a Mendelian randomization study. medRxiv 2023 doi: 10.1101/2023.06.02.23290898 [published Online First: 20230605]

46. Zhao J, Stewart ID, Baird D, et al. Causal effects of maternal circulating amino acids on offspring birthweight: a Mendelian randomisation study. (2352-3964 (Electronic))

47. Barry CA-O, Lawlor DA, Shapland CA-O, et al. Using Mendelian Randomisation to Prioritise Candidate Maternal Metabolic Traits Influencing Offspring Birthweight. LID - 10.3390/metabo12060537 [doi] LID - 537. (2218-1989 (Print))

48. Yang Q, Magnus MC, Kilpi F, et al. Investigating causal relations between sleep duration and risks of adverse pregnancy and perinatal outcomes: linear and nonlinear Mendelian randomization analyses. BMC Medicine 2022;20(1):295. doi: 10.1186/s12916-022-02494-y

49. Borges MC, Clayton GL, Freathy RM, et al. Integrating multiple lines of evidence to assess the effects of maternal BMI on pregnancy and perinatal outcomes. BMC Medicine 2024;22(1):32. doi: 10.1186/s12916-023-03167-0

50. Taylor K, McBride N, Zhao J, et al. The Relationship of Maternal Gestational Mass Spectrometry-Derived Metabolites with Offspring Congenital Heart Disease: Results from Multivariable and Mendelian Randomization Analyses. J Cardiovasc Dev Dis 2022;9(8) doi: 10.3390/jcdd9080237 [published Online First: 20220727]

51. Yang Q, Borges MC, Sanderson E, et al. Associations between insomnia and pregnancy and perinatal outcomes: Evidence from mendelian randomization and multivariable regression analyses. PLOS Medicine 2022;19(9):e1004090. doi: 10.1371/journal.pmed.1004090

52. Yang Q, Sanderson E, Tilling K, et al. Exploring and mitigating potential bias when genetic instrumental variables are associated with multiple non-exposure traits in Mendelian randomization. Eur J Epidemiol 2022;37(7):683–700. doi: 10.1007/s10654-022-00874-5 [published Online First: 20220527]

53. McBride N, Fernández-Sanlés A, Al Arab M, et al. Effects of the maternal and fetal proteome on birth weight: a Mendelian randomization analysis. medRxiv 2025 doi: 10.1101/2023.10.20.23297135 [published Online First: 20250120]

54. Brand JS, Gaillard R, West J, et al. Associations of maternal quitting, reducing, and continuing smoking during pregnancy with longitudinal fetal growth: Findings from Mendelian randomization and parental negative control studies. PLOS Medicine 2019;16(11):e1002972. doi: 10.1371/journal.pmed.1002972

