## Supplementary text for "Cohort Profile: The Mendelian Randomization in Pregnancy (MR-PREG) collaboration - Improving evidence for prevention and treatment of adverse pregnancy and perinatal outcomes"

**Contents**

Genotyping, imputation and quality control

Genome-wide association analyses

Weighted linear model (WLM)

Ethical approval

Acknowledgments

Funding

**Genotyping, imputation and quality control**

Specific genotyping, imputation and quality control (QC) procedures were applied within each cohort and are described below.

*Avon Longitudinal Study of Parents and Children (ALSPAC)*

Avon Longitudinal Study of Parents and Children (ALSPAC) children (1) were genotyped using the Illumina HumanHap550 quad chip genotyping platforms by 23andMe subcontracting the Wellcome Trust Sanger Institute, Cambridge, UK and the Laboratory Corporation of America, Burlington, NC, US. The resulting raw genome-wide data were subjected to standard QC methods. Individuals were excluded based on gender mismatches; minimal or excessive heterozygosity; disproportionate levels of individual missingness (>3%) and insufficient sample replication (IBD < 0.8). Population stratification was assessed by multidimensional scaling analysis and compared with Hapmap II (release 22) European descent (CEU), Han Chinese, Japanese and Yoruba reference populations; all individuals with non-European ancestry were removed. SNPs with a minor allele frequency of < 1%, a call rate of < 95% or evidence for violations of Hardy-Weinberg equilibrium (P < 5E-7) were removed. Cryptic relatedness was measured as proportion of identity by descent (IBD > 0.1). Related subjects that passed all other QC thresholds were retained during subsequent phasing and imputation. 9,115 subjects and 500,527 SNPs passed these QC filters.

ALSPAC mothers (2) were genotyped using the Illumina human660W-quad array at Centre National de Génotypage (CNG) and genotypes were called with Illumina GenomeStudio. PLINK (v1.07) was used to carry out QC measures on an initial set of 10,015 subjects and 557,124 directly genotyped SNPs. SNPs were removed if they displayed more than 5% missingness or a Hardy-Weinberg equilibrium P value of less than 1.0e-06. Additionally SNPs with a minor allele frequency of less than 1% were removed. Samples were excluded if they displayed more than 5% missingness, had indeterminate X chromosome heterozygosity or extreme autosomal heterozygosity. Samples showing evidence of population stratification were identified by multidimensional scaling of genome-wide identity by state pairwise distances using the four HapMap populations as a reference, and then excluded. Cryptic relatedness was assessed using a IBD estimate of more than 0.125 which is expected to correspond to roughly 12.5% alleles shared IBD or a relatedness at the first cousin level. Related subjects that passed all other QC thresholds were retained during subsequent phasing and imputation. 9,048 subjects and 526,688 SNPs passed these QC filters.

SNP genotypes in common between the sample of mothers and sample of children were combined (N = 477,482). SNPs with genotype missingness above 1% (N = 11,396 SNPs) or individuals with potential ID mismatches (N = 321 individuals) were removed. This resulted in a dataset of 17,842 subjects containing 6,305 duos and 465,740 SNPs (112 were removed during liftover and 234 were out of HWE after combination).

Haplotypes were estimated using ShapeIT (v2.r644) which utilises relatedness during phasing. The phased haplotypes were then imputed to the Haplotype Reference Consortium (HRCr1.1, 2016) panel of approximately 31,000 phased whole genomes. The HRC panel was phased using ShapeIt v2, and the imputation was performed using the Michigan imputation server. This resulted in 8,237 eligible children and 8,196 eligible mothers with available genotype data after exclusion of related subjects using cryptic relatedness measures described previously. Reference genome build version 37 (GRCh37/hg19) was used for genomic positions for ALSPAC genotype data.

*Born in Bradford (BiB)*

Born in Bradford (BiB) (3) mothers and offspring were genotyped at Bristol Bioresource Laboratories, Bristol, UK using four different Illumina arrays: HumanCoreExome12v1.0, HumanCoreExome12v1.1, HumanCoreExome24v1.0 and Infinium Global Screening Array-24 v1.0 (GSA). Genotypes were called with Illumina GenomeStudio. For each of the four subsets of participants genotyped on each array, any individual or SNP with >3% missing genotype calls was dropped and the four subsets were merged, giving a total merged sample of 20,000 individuals. It was not appropriate to apply filters for deviation from Hardy-Weinberg Equilibrium and excess homozygosity to this merged dataset, given the population structure and consanguinity known to be present. 707 individuals were then dropped due to i) mismatch between genetically- and phenotypically-defined sex (110 individuals), ii) suspected sample duplication (127 individuals), and iii) discrepancy between expected and genetically inferred relationship for first degree relatives (470 individuals). Principal component analysis (PCA) was carried out and involved assigning participants to two groups that were genetically similar to the European or South Asian HapMap samples. Ancestry data were also self-reported or obtained from primary care medical records, and categorised using the UK Office of National Statistics criteria: (1) White European ('White British' or 'White European'), (2) South Asian ('Pakistani', 'Indian' or 'Bangladeshi'), (3) Caribbean or African ('Afro-Caribbean' or 'African') or (4) others. Individuals whose PCA-based ancestry agreed with their self-reported/medical record-derived ancestry were assigned to two groups, for which we use the shorthand "White European" (WE) and "South Asian" (SA). The SA group consisted primarily of individuals with Pakistani self-reported/medical record ancestry, and does not include Bangladeshis but does include some Indians.

Genotype imputation was performed for 7606 WE and 8692 SA participants, with each ancestry and array (CoreExome/GSA) imputed separately. Prior to imputation, variants with call rate <95%, HWE P-value <1e-6 or MAF <1% were removed for each of the four ancestry/array groups separately. A/T and C/G SNPs were dropped, as were those that failed checks for strand, reference/alternative alleles, position, and SNP duplication, indels, non-autosomal SNPs, SNPs with no match in the HRC panel (position or SNP ID), and SNPs with non-matching alleles. Genotype data were imputed to the HRC r1.1 reference panel. Reference genome build version 37 (GRCh37/hg19) was used for genomic positions for BiB genotype data.

*The Norwegian Mother Father and Child Cohort Study (MoBa)*

In The Norwegian Mother Father and Child Cohort Study (MoBa) (4), genotyping of the samples was performed in seven different batches on different Illumina platforms over a period of four years at deCODE genetics, Reykjavik, Iceland (HumanCoreExome-12 v.1.1, HumanCoreExome-24 v.1.0, Global Screening Array v.1.0, HumanOmniExpress-24-v1.0, InfiniumOmniExpress-24v1.2, and GSA24-v1.0). Reference genome build version 37 (GRCh37/hg19) was used for genomic positions for MoBa genotype data. Genotypes were called in Illumina GenomeStudio v.2011.1 for the HARVEST substudy (11,490 triads) and v.2.0.3 for the remaining batches. Cluster positions were identified from samples with call rate ≥ 0.98 and GenCall score ≥ 0.15.

Pre-imputation QC was performed for each subpopulation on the SNP, individual, and family level. Subpopulations were defined using principal component (PC) analyses using 1000 Genomes phase 1 data to identify the European, Asian, and African core subpopulations (after removing SNPs with MAF < 1%, call rate < 95%, and HWE p-value < 0.001). The primary software used for the QC were PLINK 1.9 and KING 2.2.5. For the SNP-level QC, SNPs were removed if MAF < 0.5%, call rate < 95%, HWE p-value < 0.000001, discordant in duplicate pairs, associated with genotype plate and genotype batch at p-value 0.001. For individual-level QC, individuals were removed if heterozygosity outliers Fhet ± 0.2, erroneous sex assignment, known relatedness, cryptic relatedness, identity-by-decent (PI_HAT threshold of 0.15), and PC outliers both with and without 1000 Genomes. For family-level QC, families with more than 5% Mendel errors and SNPs with more than 1% of Mendel errors were removed, while other minor Mendel errors were zeroed out. Batches that were genotyped using the same array were merged (keeping only SNPs present in all batches) and the pre-imputation QC was performed on the merged batches.

Phasing and imputation were performed using the publicly available Haplotype Reference Consortium data. Phasing was performed using SHAPEIT2 with the duoHMM algorithm to incorporate the pedigree information into the haplotype estimates. IMPUTE 4 was then used to perform imputation. Dosage data was converted to best-guess (hard call) genotype data using certainty threshold 0.7, and variants with imputation quality score (INFO) <0.8 were dropped.

Post-imputation QC was then performed as for the steps outlined for pre-imputation QC, with the following changes: i) MAF filter 1%, ii) call rate <95% for SNPs, and iii) P-value for batch effects filter <5e-8. To ensure that across batch relatedness (both known and unknown) was accounted for in all analyses the three imputation batches were then merged, and the post-imputation QC steps were repeated, with the exception that a filter was applied for imputation batch effects rather than genotype batch effects.

*UK Biobank*

A full description of the genetic QC performed by the MRC-Integrative Epidemiology Unit on UK Biobank data is available elsewhere (5). The full data release contains the cohort of successfully genotyped samples (n=488,377). 49,979 individuals were genotyped using the UK BiLEVE array and 438,398 using the UK Biobank axiom array. Pre-imputation QC, phasing and imputation are described elsewhere (6). In brief, prior to phasing, multiallelic SNPs or those with MAF ≤1% were removed. Phasing of genotype data was performed using a modified version of the SHAPEIT2 algorithm (7). Genotype imputation to a reference set combining the UK10K haplotype and HRC reference panels (8) was performed using IMPUTE2 algorithms (9). The analyses presented here were restricted to autosomal variants using a graded filtering with varying imputation quality for different allele frequency ranges. Therefore, rarer genetic variants are required to have a higher imputation INFO score (Info>0.3 for MAF >3%; Info>0.6 for MAF 1-3%; Info>0.8 for MAF 0.5-1%; Info>0.9 for MAF 0.1-0.5%) with MAF and Info scores having been recalculated on an in-house derived 'European' subset.

In post-imputation QC, individuals with sex-mismatch (derived by comparing genetic sex and reported sex) or individuals with sex chromosome aneuploidy were excluded from the analysis (n=814). The sample was restricted to individuals of 'European' ancestry as defined by an in-house k-means cluster analysis performed using the first 4 principal components provided by UK Biobank in the statistical software environment R. The current analysis includes the largest cluster from this analysis (n=464,708). Estimated kinship coefficients using the KING toolset (10) identified 107,162 pairs of related individuals. An in-house algorithm was then applied to this list and preferentially removed the individuals related to the greatest number of other individuals until no related pairs remain. These individuals were excluded (n=79,448). Additionally, 2 individuals were removed due to them relating to a very large number (>200) of individuals. Reference genome build version 37 (GRCh37/hg19) was used for genomic positions for UKB genotype data.

*FinnGen*

A full description of the FinnGen study and the QC procedures applied to the genetic data is available elsewhere (11, 12). We used release 12 of the publicly available FinnGen summary statistics. FinnGen individuals were genotyped with Illumina and Affymetrix chip arrays (Illumina Inc., San Diego, and Thermo Fisher Scientific, Santa Clara, CA, USA). Genotype calls were made with GenCall and zCall algorithms for Illumina and AxiomGT1 algorithm for Affymetrix data. Chip genotyping data produced with previous chip platforms and reference genome builds were lifted over to build version 38 (GRCh38/hg38). In sample-wise QC steps, individuals with ambiguous gender, high genotype missingness (>5%), excess heterozygosity (+-4SD) and non-Finnish ancestry were excluded. In variant-wise QC steps, variants with high missingness (>2%), low HWE P-value (<1e-6) and low minor allele count (MAC<3) were excluded. Before imputation, chip-genotyped samples were pre-phased with Eagle 2.3.5 using the default parameters, except the number of conditioning haplotypes, which was set to 20,000. Chip genotype data were imputed using the population-specific SISu v4.2 imputation reference panel of 8,554 whole genomes with Beagle 4.1 (version 27Jan18.7e1). Post-imputation QC involved checking the expected conformity of the imputation INFO-value distribution, MAF differences between the target dataset and the imputation reference panel and checking chromosomal continuity of the imputed genotype calls. Merged imputed genotype data is composed of 141 data sets and includes samples from multiple cohorts, yielding a total sample size of 520,210 participants with genotype data for 21,311,644 variants.

The FinnGen data for 180,042 pruned SNPs was merged with the 1k genome project (1kgp) data, and two rounds of PCA were carried out. After kinship analysis, the sample was separated into three groups: 296,829 inliers (unrelated participants with Finnish ancestry), 203,908 outliers (non-duplicate samples with Finnish ancestry but who are also related to the inliers), and 19,473 rejected participants who had non-Finnish ancestry, or who were twins/duplicates with relations to other (non-inlier) samples. A final round of PCA for the inliers was calculated, after which outliers were projected onto the same PC space, yielding PC covariates for a total of 500,737 participants. After dropping 355 participants due to missing minimum phenotype data, and dropping 34 participants due to sex check failure, 500,348 participants were available for genome-wide association study (GWAS) analyses, of which 282,064 were female. Reference genome build version 38 (GRCh38/hg38) was used for genomic positions for FinnGen.

### Genome-wide association analyses

Genome-wide association study (GWAS) analyses were run separately for mothers, fathers and offspring (when sufficient data were available) within each cohort, using software and approaches which were tailored to optimally account for the population stratification and relatedness present in each sample.

*ALSPAC*

We performed GWAS analysis in ALSPAC mothers and offspring using PLINK 2.0 alpha (build 22 Dec 2024) (13, 14). The analytical samples were restricted to unrelated mothers (IBD ≤ 0.125) and offspring (IBD ≤ 0.1). Association analyses were performed with imputed autosomal genotype data, for adverse pregnancy and perinatal outcomes (APPOs) via the "--glm" command using Firth logistic regression for binary outcomes and linear regression for continuous outcomes. Firth regression enables less biased effect size estimation in the presence of case-control imbalance for binary outcomes. We assumed an additive model and adjusted for the top 10 principal components of ancestry (PCs). PCs were generated using Plink –pca option after removing related individuals based on a set of independent SNPs excluding any long-range LD regions (variant count window size = 100, variant count to shift the window = 5, variance inflation factor (VIF) threshold = 1.01).

*BiB*

We performed GWAS analysis in BiB mothers and offspring using REGENIE version 3.6 (15) to account for the complex population structure (population stratification and relatedness) that is present in BiB, via a whole-genome regression approach. REGENIE implements whole-genome regression in two steps: in step 1, array genotypes are used to generate 22 leave-one-chromosome-out (LOCO) predictions for each outcome. In step 2, during which the imputed genotype associations with each outcome are estimated, these LOCO predictions are then used as covariates. We performed GWAS separately for the two largest ancestry groups present in BiB as described above (SA and WE). We did not use REGENIE's automatic phenotype imputation feature for missing APPOs and instead ran REGENIE for each outcome separately. For step 1 we used the called genotype data obtained by merging the samples genotyped on different arrays (as described above), after which the PLINK 2.0 filters "--maf 0.01", "--hwe 0.000001" and "--geno 0.03" were applied, yielding sets of 99,131 and 135,801 variants for the SA and WE samples respectively. For step 1 we set genotype block size ("--bsize") to 1000. For step 2, we ran association analyses with imputed autosomal genotype data, having first merged the samples genotyped on the CoreExome and GSA arrays and subsetted variants to those with imputation accuracy $R^2$ ≥0.3 when imputed from both arrays. We set genotype block size to 200, applied a minimum minor allele count (MAC) filter of 20, and used linear regression or Firth logistic regression (via REGENIE options "--firth --approx --pThresh 0.999999") for continuous and binary outcomes respectively. We computed the standard error based on the Firth regression effect size and likelihood ratio test *P*-value as per the "--firth-se" option. Models were adjusted for genotyping array and 40 PCs: 20 internal PCs (calculated using only BiB data via the flashpca package version 2.0 (16) with the authors' recommended settings (16, 17)) and 20 external PCs (BiB participants were projected onto a UK Biobank PC space via the pcapred R package (18).

*MoBa*

We performed GWAS analysis in MoBa mothers, fathers and offspring using whole genome regression implemented in REGENIE version 3.1.2 (15). We did not use REGENIE's automatic phenotype imputation feature for missing APPOs and instead ran REGENIE for each outcome separately. We performed step 1 using 455,827 imputed genetic variants which were directly genotyped in ≥1 imputation batch and had imputation quality INFO score >0.99785, having first removed 35,810 variants in the major histocompatibility complex (MHC) region on chromosome 6 (variants with MAF <1% had already been removed in pre-phase QC). We set genotype block size ("--bsize") to 1000. For step 2, we ran association analyses with 6,981,748 imputed (hard called) autosomal genotype data, adjusting for genotyping batch and 20 PCs. We set genotype block size to 200 and used linear regression or Firth logistic regression (via REGENIE options "--firth --approx --pThresh 0.999999") for continuous and binary outcomes respectively. We computed the standard error based on the Firth regression effect size and likelihood ratio test *P*-value as per the "--firth-se" option.

*UK BioBank*

We performed GWAS analysis in UK Biobank mothers only, using whole genome regression implemented in REGENIE version v3.6. (15). We did not use REGENIE's automatic phenotype imputation feature for missing APPOs and instead ran REGENIE for each outcome separately. We performed step 1 using 549,043 directly genotyped variants with MAF ≥0.01, variant call rate ≥0.9; Hardy-Weinberg equilibrium *P* >1.0 x $10^{-15}$ and MAC ≥100, and set genotype block size ("--bsize") to 1000. For step 2, we ran association analyses with imputed genotype data for the APPOs, adjusting for genotyping array and 40 PCs. We set genotype block size to 200 and used linear regression or Firth logistic regression (via REGENIE options "--firth --approx --pThresh 0.999999") for continuous and binary outcomes respectively. We computed the standard error based on the Firth regression effect size and likelihood ratio test *P*-value as per the "--firth-se" option.

*FinnGen*

GWAS analyses were caried out in FinnGen mothers only, using whole -genome regression implemented in REGENIE version 2.2.4 (15). For REGENIE step 1, 215,152 pruned (1.5Mb window and r2 threshold of 0.2) variants were used after applying the following filters: i) imputation INFO score >0.95 in all batches, ii) >97 % non-missing genotype, and iii) MAF >1 %, and a genotype block size of 1000 was used. For REGENIE step 2, association analyses with imputed genotype data for APPOs were conducted for each variant with a minimum allele count of 5 among each phenotype's cases and controls. The approximate Firth test was used for variants with an initial *P*-value of less than 0.01 and the standard error was computed based on the Firth regression effect size and likelihood ratio test *P*-value (REGENIE options --firth --approx --pThresh 0.01 --firth-se).

***Post-GWAS quality control***

Prior to meta-analysis we excluded variants with low MAF (<0.01) or low INFO score (<0.4) from the summary statistics from individual cohorts and maternal/paternal/offspring samples. We applied a standardised QC pipeline to GWAS summary statistics from individual cohorts and from the meta-analysis using the GWASInspector R package (19). This QC process included comparing the allele frequencies (AF) in the GWAS summary statistics to those in suitable reference panels. In the FinnGen summary statistics a small subset of variants had the opposite AF to the HRC reference panel (i.e. $AF_{FinnGen} = 1 - AF_{HRC}$). We therefore dropped 4064 variants which had a FinnGen versus HRC AF difference >0.2 from the meta-analyses.

***Weighted linear model (WLM)***

We carried out marginal (unadjusted) GWAS as described above, in which outcomes were regressed separately on maternal and offspring genotype, without mutual adjustment of the effects of maternal and offspring genotypes. After meta-analysis of the marginal GWAS effects we applied a weighted linear model (WLM) (21-23) implemented in the DONUTS R package (24) to estimate conditional (adjusted) maternal ($\hat{\beta}_m$) and offspring ($\hat{\beta}_o$) genetic effects (the mutually adjusted coefficients for maternal and offspring genotype, fitted jointly in the same model), as linear combinations of the estimated marginal offspring ($\hat{b}_o$) and maternal ($\hat{b}_m$) genetic effects:

$$\hat{\beta}_o = \frac{4}{3}\hat{b}_o - \frac{2}{3}\hat{b}_m$$

$$\hat{\beta}_m = \frac{4}{3}\hat{b}_m - \frac{2}{3}\hat{b}_o$$

$$\mathrm{SE}(\hat{\beta}_o) = \sqrt{\frac{16}{9}\mathrm{var}(\hat{b}_o) + \frac{4}{9}\mathrm{var}(\hat{b}_m) - \frac{16}{9} \times \widehat{\mathrm{int}}_{o,m} \times \mathrm{SE}(\hat{b}_m)\mathrm{SE}(\hat{b}_o)}$$

$$\mathrm{SE}(\hat{\beta}_m) = \sqrt{\frac{16}{9}\mathrm{var}(\hat{b}_m) + \frac{4}{9}\mathrm{var}(\hat{b}_o) - \frac{16}{9} \times \widehat{\mathrm{int}}_{o,m} \times \mathrm{SE}(\hat{b}_m)\mathrm{SE}(\hat{b}_o)}$$

where $\widehat{\mathrm{int}}_{o,m}$ is the intercept term from bivariate LD score regression (25) of the maternal and offspring marginal summary statistics. These intercept terms are necessary to account for sample overlap between the marginal GWAS samples, and were estimated using Hapmap 3 SNPs with MAF >1%, excluding SNPs in the MHC region and with association Chi square statistics >80. We refer to the above model as the duos WLM, and it assumes that conditional paternal effects are zero.

Paternal genotype data were available in MoBa. We therefore also conducted marginal paternal GWAS, enabling us to relax the assumption that conditional paternal effects are zero by estimating the conditional maternal, offspring and paternal ($\hat{\beta}_p$) genetic effects via a trios WLM:

$$\hat{\beta}_o = 2\hat{b}_o - \hat{b}_m - \hat{b}_p$$

$$\hat{\beta}_m = \frac{3}{2}\hat{b}_m - \hat{b}_o + \frac{1}{2}\hat{b}_p$$

$$\hat{\beta}_p = \frac{3}{2}\hat{b}_p - \hat{b}_o + \frac{1}{2}\hat{b}_m$$

with their standard errors:

$$\mathrm{SE}(\hat{\beta}_o) = \sqrt{4\mathrm{var}(\hat{b}_o) + \mathrm{var}(\hat{b}_m) + \mathrm{var}(\hat{b}_p) + 2 \times \mathrm{int}_{m,p} \times \mathrm{SE}(\hat{b}_m)\mathrm{SE}(\hat{b}_p) - 4 \times \mathrm{int}_{o,m} \times \mathrm{SE}(\hat{b}_o)\mathrm{SE}(\hat{b}_m) - 4 \times \mathrm{int}_{o,p} \times \mathrm{SE}(\hat{b}_o)\mathrm{SE}(\hat{b}_p)}$$

$$\mathrm{SE}(\hat{\beta}_m) = \sqrt{\frac{9}{4}\mathrm{var}(\hat{b}_m) + \mathrm{var}(\hat{b}_o) + \frac{1}{4}\mathrm{var}(\hat{b}_p) - \mathrm{int}_{o,p} \times \mathrm{SE}(\hat{b}_o)\mathrm{SE}(\hat{b}_p) - 3 \times \mathrm{int}_{o,m} \times \mathrm{SE}(\hat{b}_o)\mathrm{SE}(\hat{b}_m) + \frac{3}{2} \times \mathrm{int}_{m,p} \times \mathrm{SE}(\hat{b}_m)\mathrm{SE}(\hat{b}_p)}$$

$$\mathrm{SE}(\hat{\beta}_p) = \sqrt{\frac{9}{4}\mathrm{var}(\hat{b}_p) + \mathrm{var}(\hat{b}_o) + \frac{1}{4}\mathrm{var}(\hat{b}_m) - \mathrm{int}_{o,m} \times \mathrm{SE}(\hat{b}_o)\mathrm{SE}(\hat{b}_m) - 3 \times \mathrm{int}_{o,p} \times \mathrm{SE}(\hat{b}_o)\mathrm{SE}(\hat{b}_p) + \frac{3}{2} \times \mathrm{int}_{m,p} \times \mathrm{SE}(\hat{b}_m)\mathrm{SE}(\hat{b}_p)},$$

where $\mathrm{int}_{o,m}$, $\mathrm{int}_{o,p}$, and $\mathrm{int}_{m,p}$ are the intercepts from bivariate LD score regression of the maternal and offspring, paternal and offspring and maternal and paternal marginal summary statistics respectively, and $\hat{b}_p$ is the estimated marginal paternal genetic effect.

The DONUTS software package extends the duos and trios WLMs to account for assortative mating via a parameter α. We set α to zero (i.e. we assumed mating was random) because previous sensitivity analyses in MoBa suggested this gave the most accurate estimates for conditional effects on birth weight [Bond et al medrXiv].

**Ethical approval**

*ALSPAC*

Ethical approval for the study was obtained from the ALSPAC Ethics and Law Committee and the Local Research Ethics Committees. Consent for biological samples has been collected in accordance with the Human Tissue Act (2004).

Ethical approval was obtained from the ALSPAC Ethics and Law Committee and the Local Research Ethics Committees. Consent for biological samples has been collected in accordance with the Human Tissue Act (2004). Informed consent for the use of data collected via questionnaires and clinics was obtained from participants following the recommendations of the ALSPAC Ethics and Law Committee at the time (details and reference numbers of all ethics approvals can be found at http://www.bristol.ac.uk/media-library/sites/alspac/documents/governance/Research%20Ethics%20Committee%20approval%20references.pdf).

*BiB*

Ethical approval for the study was granted by the Bradford National Health Service Research Ethics Committee (ref 06/Q1202/48), and all participants gave written informed consent. The ALL IN sub-study had ethical approval from the London School of Hygiene & Tropical Medicine ethics committee (ref: 5320) and the Bradford Research Ethics committee (ref: 08/H1302/21). Parents (usually the mother) gave informed, written consent to take part in the study.

*MoBa*

The current study is based on version 12 of the quality-assured data files released for research in 2019. The establishment of MoBa and initial data collection was based on a license from the Norwegian Data Protection Agency and approval from The Regional Committees for Medical and Health Research Ethics. The MoBa cohort is currently regulated by the Norwegian Health Registry Act. The current study was approved by The Regional Committees for Medical and Health Research Ethics of South/East Norway (ref 2018/1256).

*UK Biobank*

The UK Biobank has approval from the North West Multi-centre Research Ethics Committee (MREC) as a Research Tissue Bank (RTB) approval. This RTB approval was granted initially in 2011 (11/NW/0382) and it is renewed on a 5-yearly cycle, with the latest one successfully renewed in 2021 (21/NW/0157).

**Acknowledgments**

*ALSPAC*

We are extremely grateful to all the families who took part in this study, the midwives for their help in recruiting them, and the whole ALSPAC team, which includes interviewers, computer and laboratory technicians, clerical workers, research scientists, volunteers, managers, receptionists and nurses.

*BiB*

BiB is only possible because of the enthusiasm and commitment of the Children and Parents in BiB. We are grateful to all the participants, teachers, school staff, health professionals and researchers who have made BiB happen.

*MoBa*

This research has been conducted using MoBa data using application number 2552. MoBa is supported by the Norwegian Ministry of Health and Care services and the Ministry of Education and Research. We are grateful to all the participating families in Norway who take part in this on-going cohort study. We thank the Norwegian Institute of Public Health (NIPH) for generating high-quality genomic data. This research is part of the HARVEST collaboration, supported by the Research Council of Norway (#229624). We also thank the NORMENT Centre for providing genotype data, funded by the Research Council of Norway (#223273), South East Norway Health Authority and KG Jebsen Stiftelsen. We further thank the Center for Diabetes Research, the University of Bergen for providing genotype data and performing QC and imputation of the data funded by the ERC AdG project SELECTionPREDISPOSED, Stiftelsen Kristian Gerhard Jebsen, Trond Mohn Foundation, the Research Council of Norway, the Novo Nordisk Foundation, the University of Bergen, and the Western Norway health Authorities (Helse Vest).

*FinnGen*

The authors thank the FinnGen investigators for sharing their summary-level data.

*UK Biobank*

We would like to thank all the participants of UK Biobank for their vital contribution to the resource. This research has been conducted using the UK Biobank Resource under Application Number 23938.

**Funding**

*ALSPAC*

The UK Medical Research Council and Wellcome (Grant ref: 217065/Z/19/Z) and the University of Bristol provide core support for ALSPAC.A comprehensive list of grants funding is available on the ALSPAC website (http://www.bristol.ac.uk/alspac/external/documents/grant-acknowledgements.pdf). ALSPAC GWAS data was generated by Sample Logistics and Genotyping Facilities at Wellcome Sanger Institute and LabCorp (Laboratory Corporation of America) using support from 23andMe. This research was funded in part by the Wellcome Trust (Grant ref: 217065/Z/19/Z)]. For the purpose of Open Access, the author has applied a CC BY public copyright licence to any Author Accepted Manuscript version arising from this submission.

*MoBa*

MoBa funding is under **Acknowledgements** as requested by MoBa publication guidelines.

*BiB*

BiB receives core funding from the Wellcome Trust (WT101597MA), a joint grant from the UK Medical and Economic and Social Science Research Councils (MR/N024397/1), British Heart Foundation (CS/16/4/32482), and the National Institute of Health Research under its Applied Research Collaboration for Yorkshire and Humber and Clinical Research Network research delivery support. Further support for genome-wide and multiple 'omics measurements in BiB is from the UK Medical Research Council (G0600705), National Institute of Health Research (NF-SI-0611-10196), US National Institute of Health (R01DK10324), and the European Research Council under the European Union's Seventh Framework Programme (FP7/2007–2013) / ERC grant agreement no 669545.

*UK Biobank*

UK Biobank is funded primarily by the Wellcome Trust and the Medical Research Council (MRC). It is also funded by the Department of Health, British Heart Foundation, Cancer Research UK, Diabetes UK, National Institute for Health Research (NIHR), Scottish Government, Northwest Regional Development Agency, and Welsh Assembly Government.

### References


1. Boyd A, Golding J, Macleod J, Lawlor DA, Fraser A, Henderson J, et al. Cohort profile: the 'children of the 90s'—the index offspring of the Avon Longitudinal Study of Parents and Children. Int J Epidemiol. 2013;42(1):111-27.
2. Fraser A, Macdonald-Wallis C, Tilling K, Boyd A, Golding J, Davey Smith G. Cohort profile: The Avon Longitudinal Study of Parents and Children: ALSPAC mothers cohort. Int J Epidemiol. 2013;42.
3. Wright J, on behalf of the Born in Bradford Scientific Collaborators G, Small N, Raynor P, Tuffnell D, Bhopal R, et al. Cohort Profile: The Born in Bradford multi-ethnic family cohort study. International Journal of Epidemiology. 2013;42(4):978-91.
4. Corfield EC, Frei O, Shadrin AA, Rahman Z, Lin A, Athanasiu L, et al. The Norwegian Mother, Father, and Child cohort study (MoBa) genotyping data resource: MoBaPsychGen pipeline v.1. bioRxiv. 2022:2022.06.23.496289.
5. Mitchell R, Hemani G, Dudding T, Corbin L, Harrison S, Paternoster L. UK Biobank Genetic Data: MRC-IEU Quality Control, version 2. 2019.
6. Bycroft C, Freeman C, Petkova D, Band G, Elliott LT, Sharp K, et al. The UK Biobank resource with deep phenotyping and genomic data. Nature. 2018;562(7726):203-9.
7. O'Connell J, Sharp K, Shrine N, Wain L, Hall I, Tobin M, et al. Haplotype estimation for biobank-scale data sets. Nat Genet. 2016;48(7):817.
8. Huang J, Howie B, McCarthy S, Memari Y, Walter K, Min JL, et al. Improved imputation of low-frequency and rare variants using the UK10K haplotype reference panel. Nature Communications. 2015;6:8111.
9. Howie B, Marchini J, Stephens M. Genotype Imputation with Thousands of Genomes. G3 Genes|Genomes|Genetics. 2011;1(6):457-70.
10. Manichaikul A, Mychaleckyj JC, Rich SS, Daly K, Sale M, Chen W-M. Robust relationship inference in genome-wide association studies. Bioinformatics. 2010;26(22):2867-73.
11. Kurki MI, Karjalainen J, Palta P, Sipilä TP, Kristiansson K, Donner KM, et al. FinnGen provides genetic insights from a well-phenotyped isolated population. Nature. 2023;613(7944):508-18.
12. FinnGen. Documentation of R12 release 2024 [Available from: https://finngen.gitbook.io/documentation/.
13. Purcell S, Chang C. PLINK. 2.00 alpha (build 22 Dec 2024) ed2024.
14. Chang CC, Chow CC, Tellier LC, Vattikuti S, Purcell SM, Lee JJ. Second-generation PLINK: rising to the challenge of larger and richer datasets. Gigascience. 2015;4(1):7.
15. Mbatchou J, Barnard L, Backman J, Marcketta A, Kosmicki JA, Ziyatdinov A, et al. Computationally efficient whole-genome regression for quantitative and binary traits. Nature Genetics. 2021;53(7):1097-103.
16. Abraham G, Qiu Y, Inouye M. FlashPCA2: principal component analysis of Biobank-scale genotype datasets. Bioinformatics. 2017;33(17):2776-8.
17. Abraham G. FlashPCA2 2016 [Available from: https://github.com/gabraham/flashpca.
18. Lawson D. R package pcapred 2020 [Available from: https://github.com/danjlawson/pcapred.
19. Ani A, van der Most PJ, Snieder H, Vaez A, Nolte IM. GWASinspector: comprehensive quality control of genome-wide association study results. Bioinformatics. 2021;37(1):129-30.
20. Willer CJ, Li Y, Abecasis GR. METAL: fast and efficient meta-analysis of genomewide association scans. Bioinformatics. 2010;26(17):2190-1.
21. Beaumont RN, Flatley C, Vaudel M, Wu X, Chen J, Moen G-H, et al. Genome-wide association study of placental weight in 179,025 children and parents reveals distinct and shared genetic influences between placental and fetal growth. medRxiv. 2022:2022.11.25.22282723.

22. Warrington NM, Beaumont RN, Horikoshi M, Day FR, Helgeland Ø, Laurin C, et al. Maternal and fetal genetic effects on birth weight and their relevance to cardio-metabolic risk factors. Nat Genet. 2019:1.
23. Warrington NM, Hwang L-D, Nivard MG, Evans DM. Estimating direct and indirect genetic effects on offspring phenotypes using genome-wide summary results data. Nature Communications. 2021;12(1):5420.
24. Wu Y, Zhong X, Lin Y, Zhao Z, Chen J, Zheng B, et al. Estimating genetic nurture with summary statistics of multigenerational genome-wide association studies. Proceedings of the National Academy of Sciences. 2021;118(25):e2023184118.
25. Bulik-Sullivan B, Finucane HK, Anttila V, Gusev A, Day FR, Loh P-R, et al. An atlas of genetic correlations across human diseases and traits. Nat Genet. 2015;47(11):1236-41.
