## Supplementary figures for "Cohort Profile: The Mendelian Randomization in Pregnancy (MR-PREG) collaboration - Improving evidence for prevention and treatment of adverse pregnancy and perinatal outcomes"

Avon Longitudinal Study of Parents and Children

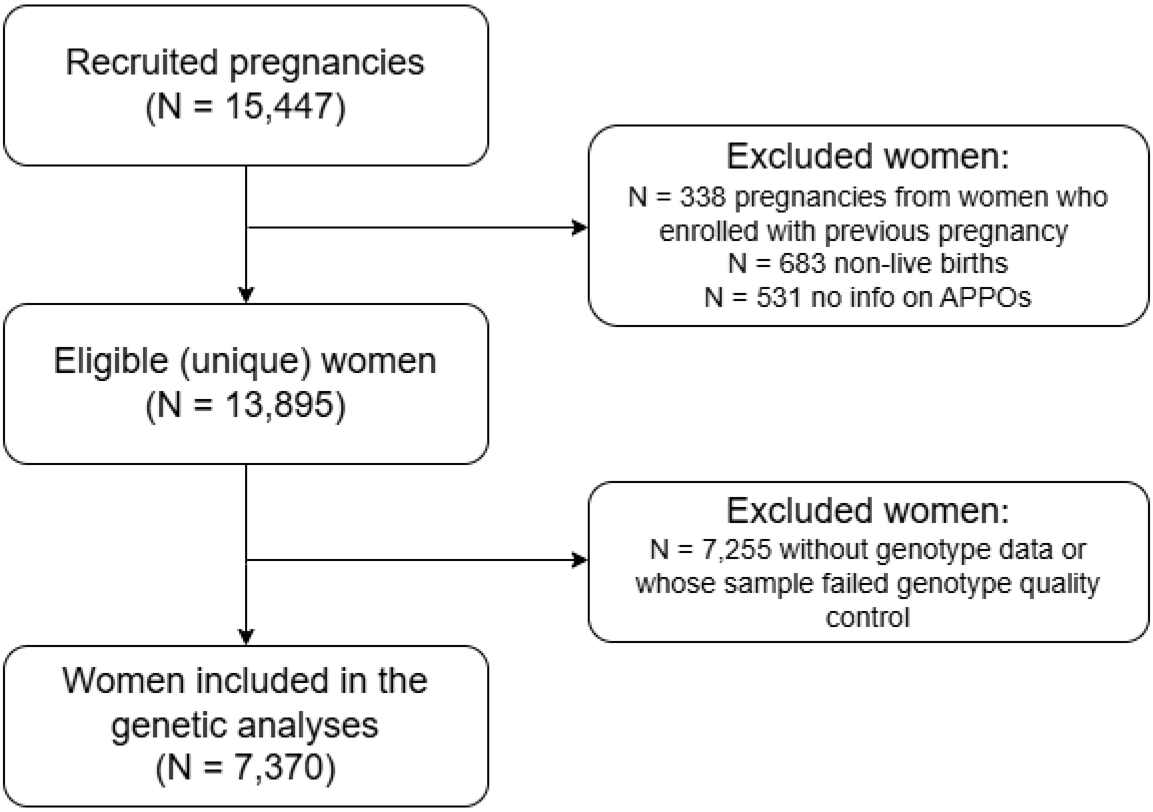


Genetic data is available for a subsample of mothers

Born in Bradford

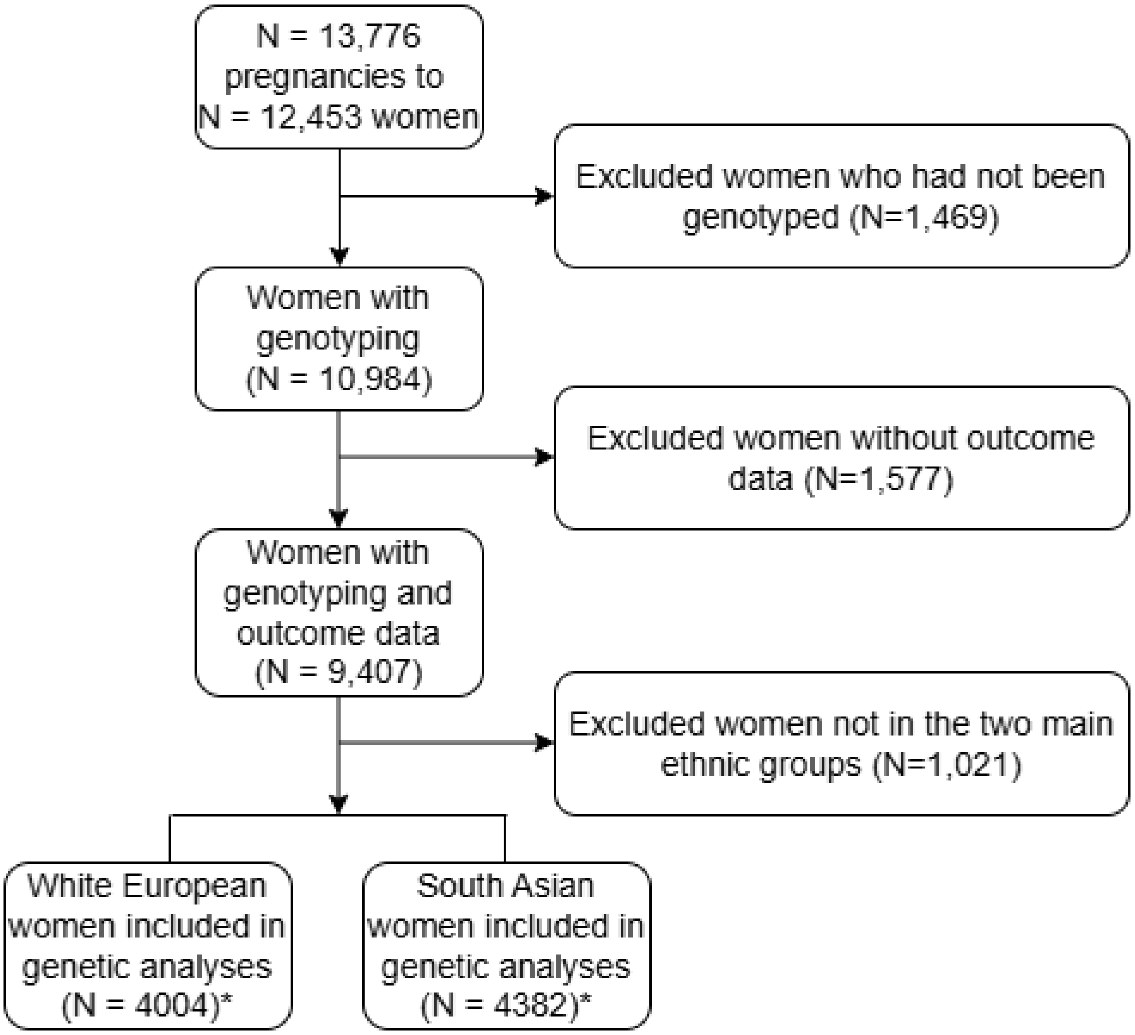


*analysed separately

Norwegian Mother Father and Child birth cohort

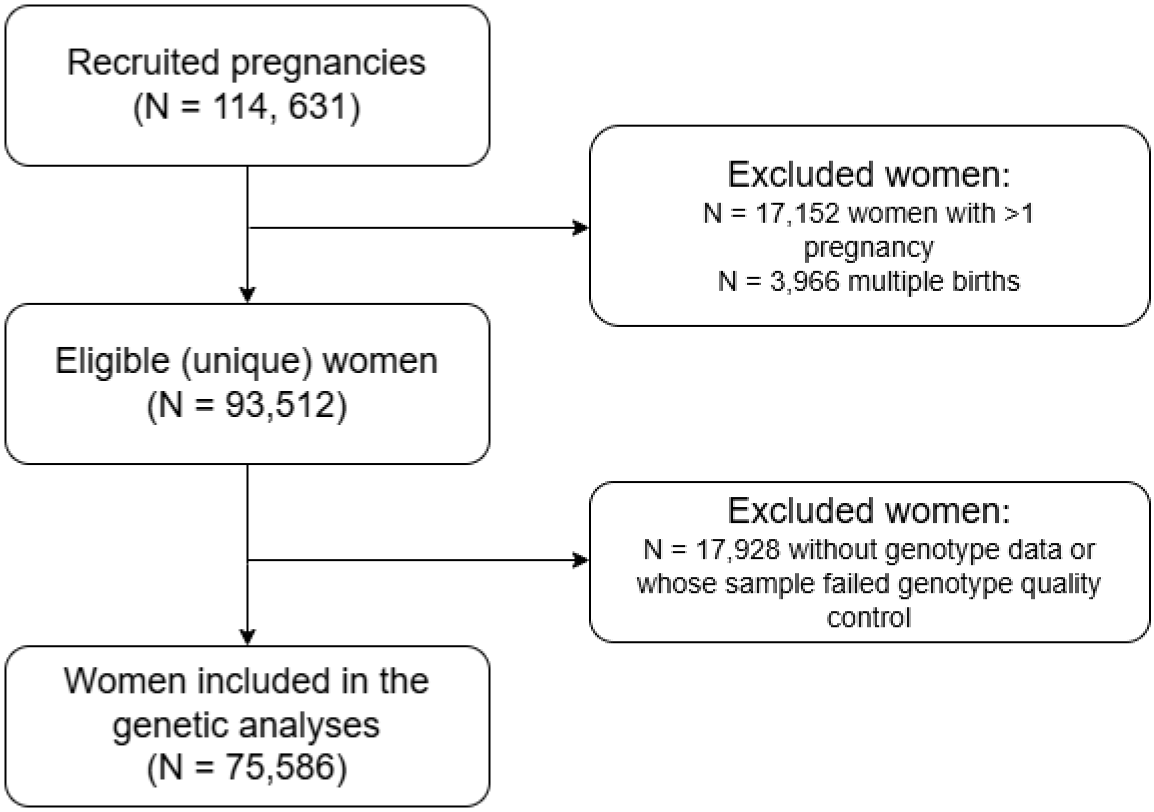

UK Biobank

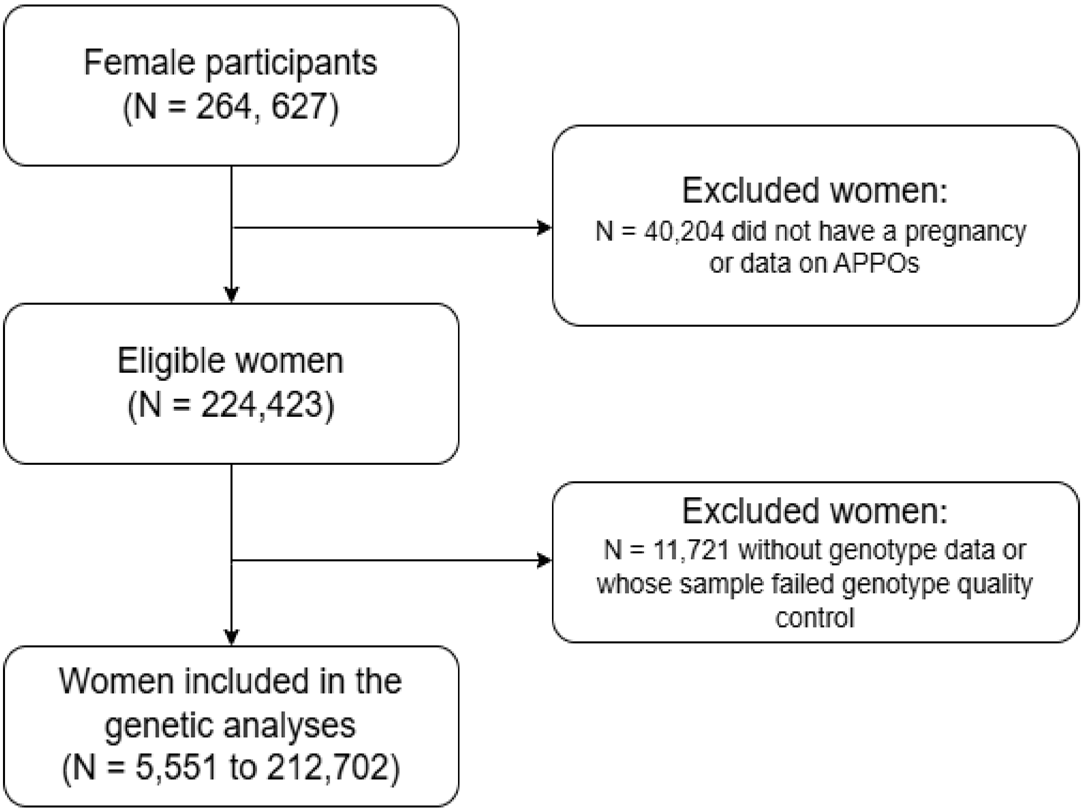


Note: some phenotypes have much smaller sample size because the source of information was maternity records or follow-up questionnaires, and these were not available for all women
